# Learning shared forecast-error structure to improve ensemble forecasts of seasonal respiratory outbreaks

**DOI:** 10.64898/2026.07.20.26358508

**Authors:** Yuchen Qin, Hongru Du, Sen Pei

**Affiliations:** Department of Systems & Information Engineering, University of Virginia, Charlottesville, VA, USA; Department of Environmental Health Sciences, Mailman School of Public Health, Columbia University,New York, NY,USA

**Keywords:** ensemble forecasts, influenza forecasting, residual learning, error correction

## Abstract

Real-time forecasts of seasonal respiratory outbreaks are critical for public health preparedness and healthcare planning. Multi-model ensembles, which combine predictions from individual models, have become a leading approach for operational outbreak forecasting. Their success, however, depends in part on the assumption that component models make sufficiently independent errors. Here, we examined this assumption using archived real-time forecasts for influenza hospitalizations and influenza-like illness (ILI) in the United States. We found that component models with diverse structures and calibration methods shared systematic forecast errors during epidemic growth and around epidemic peaks, reflecting the common challenge of tracking rapid changes in epidemic dynamics from real-time surveillance data. Because such shared errors cannot be fully corrected by ensembling alone, we developed a deep learning framework that learns structured residual errors from historical forecasts and uses them to correct ensemble predictions. This framework improved influenza hospitalization forecasts across horizons and geographic scales, reducing the Weighted Interval Score by up to 20% at the national level and 12% across states relative to official ensemble forecasts, with the largest improvements at the near-term horizon and during epidemic growth and peak periods. We further showed that learned residual structures transferred across ensembles formed from different component models, making the approach robust to changes in model participation across seasons. The framework also improved ensemble forecasts for ILI, although gains were more modest. These findings reveal a fundamental challenge in ensemble forecasting and provide a generalizable approach for improving real-time epidemic forecasts.

## Introduction

Real-time forecasting of seasonal respiratory epidemics is important for public health preparedness and healthcare response. Accurate short-term forecasts can support time-sensitive decisions about resource allocation, hospital operations, and public communication [1]. These needs are most acute during rapid epidemic growth and near peaks, when healthcare systems face surging demand and public health officials must make timely decisions about interventions and resource deployment [2]. Improving the reliability of real-time influenza forecasts during these critical periods remains a key challenge in operational epidemic prediction.

Over the past decade, influenza forecasting has become one of the most mature settings for real-time epidemic prediction, supported by repeated multi-season forecasting challenges across different targets, horizons, and spatial locations [2–4]. Among the many approaches developed, ensemble forecasting has emerged as the most consistently effective strategy [5]. By combining predictions from multiple models with different structural assumptions and inductive biases, ensemble methods reduce forecast variance, improve calibration, and provide more stable performance across seasons and locations [2, 6, 7]. However, important limitations remain. Even well-constructed ensemble forecasts can miss rapid trend changes, overshoot epidemic peaks, and lose reliability around inflection points [8], the periods when accurate forecasts carry the greatest value for public health decision-making.

Most efforts to improve epidemic forecasts have focused on the forecast generation process itself. These include developing better models (e.g., mechanistic [9, 10], statistical [11], and machine learning [12]), as well as incorporating additional data streams, such as internet search queries [13–15], viral component data [16–18], mobility data [19–21], and wastewater surveillance [22], to strengthen available signals. At the ensemble level, substantial effort has been devoted to refining how forecasting models are combined. Approaches range from simple equal-weight averages [3, 23] to trained weighting strategies based on historical performance [24–26], adaptive methods that adjust weights over time [27–30], density combination through linear and beta-transformed pooling [31], and phase-informed ensemble strategies that adjust model contributions based on epidemic stage [32].

Recent work has also examined ensemble composition, investigating the minimum number of component models needed for robust accuracy [33], whether selecting for higher-performing subsets improves upon equal-weight inclusion, and whether model diversity meaningfully affects aggregate performance [34]. Yet, despite this methodological diversity, more sophisticated weighting strategies have generally struggled to consistently outperform simple equal-weight ensembles, as individual models’ performance varies across time periods, epidemic phases, and geographic locations [23, 27].

While previous research has substantially advanced ensemble forecasting, less attention has been paid to formally quantifying the structure of forecast errors across component models. By design, ensemble approaches improve predictions by averaging errors from different models. For example, if individual model errors are independent and unbiased with distribution *N* (0, *σ*^2^), the standard deviation of the equal-weight ensemble error decreases as 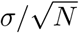, where *N* is the number of component models and *σ*^2^ is the error variance of each model. However, if individual model errors are correlated with average pairwise correlation *ρ*, the standard deviation converges to 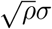 as *N* increases (*Supporting Information, Ensemble errors for individual models with correlated errors*). Thus, ensemble errors do not continue to shrink with additional component models when errors are shared or structurally correlated. Characterizing these shared forecast-error patterns is therefore critical to understanding the limits of ensemble forecasting and improving its performance.

Here, we hypothesize that real-time influenza forecast errors contain predictable seasonal, temporal, and spatial structure that can be exploited to improve prediction. We developed a Seasonal– Temporal–Spatial Residual Learning framework (STS-Residual) that operates as a model-correction procedure by learning systematic forecast errors. The framework decomposes forecast error into three complementary components: (1) a seasonal component that captures recurring epiweek-specific error patterns, (2) a temporal component that captures recent local error dynamics, and (3) a spatial component that captures cross-location dependence in forecast errors. These components are adaptively weighted to generate a real-time correction, which is then applied to the original ensemble prediction.

We evaluated this framework using archived real-time forecasts of influenza hospitalizations and influenza-like illness (ILI) in the United States (U.S.) [2, 3, 5]. STS-Residual improved short-term hospitalization forecasts across horizons and geographic scales, reducing Weighted Interval Score (WIS) by 11.6%, 6.9%, 5.0%, and 2.7% (averaged across all states) for 0-to 3-week-ahead hospitalization forecasts relative to the official FluSight ensemble. For comparison, ILI forecasts exhibited a weaker residual structure and showed more modest gains, with improvements concentrated at the nowcast horizon (6.1%) and declining to nearly neutral at longer lead times. These results suggest that shared forecast errors are operationally learnable when the residual structure is sufficiently strong and that residual learning can serve as a practical correction layer for ensemble forecasts.

## Results

### Shared error structure suggests learnable forecast residuals

We examined whether forecast errors exhibited shared structure across models and seasons using two datasets of archived real-time forecasts for seasonal respiratory outbreaks: (1) FluSight influenza hospitalization forecasts from the 2023–2024 through 2025–2026 seasons at the national and state levels, including the 50 U.S. states, the District of Columbia, and Puerto Rico; and (2) FluSight influenza-like illness (ILI) forecasts from the 2010–2011 through 2016–2017 seasons at the national level and across the 10 HHS regions. We focused on short-term predictions, defined as *h*-week-ahead forecasts for *h* = 0, 1, 2, 3.

We first analyzed influenza hospitalization forecasts from 12 models without missing predictions, including models submitted by individual teams and the official FluSight ensemble (see detailed inclusion criteria in *Supporting Information, Retained forecast models*, Tables S1–S2). For 0-week-ahead prediction, although individual model errors varied in magnitude, their trajectories were highly correlated within each season (Fig. 1A-C). In all three seasons, the mean error among the 12 models exhibited a consistent epidemic-phase-dependent pattern: models tended to underestimate hospitalizations during periods of rapid epidemic growth and to overestimate shortly after turning points.

**Figure 1:**
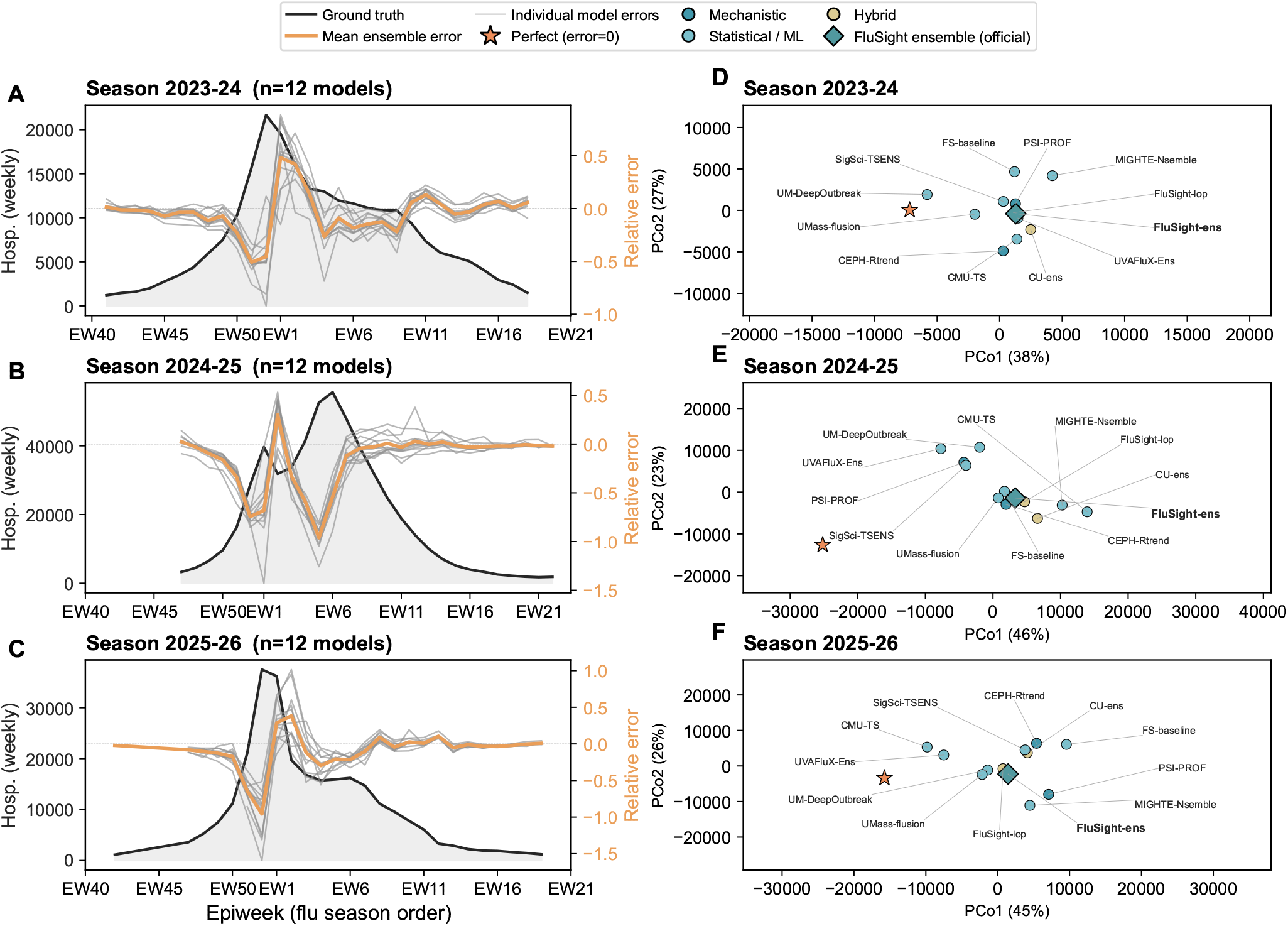
Structural errors in real-time forecasts for influenza hospitalizations. (A-C) 0-week-ahead forecast errors of component models (gray solid lines) and the unweighted ensemble (orange solid lines), overlaid with the weekly hospitalizations at the national level (black solid lines). Forecast errors are expressed as relative errors, each normalized by the season-mean target value (right axis). We include 12 models with no missing forecasts submitted to the CDC FluSight challenge (including the FluSight ensemble). For the 2025-2026 season, data are shown up to Epiweek 18 in 2026, when the analysis was performed. (D-F) Two-dimensional Principal Coordinates Analysis (PCoA, classical metric multidimensional scaling) of 0-week-ahead forecast-error time series, computed from pairwise Euclidean distances, with a synthetic ‘Perfect’ (zero-error) reference included for context. PCo1 and PCo2 together explain approximately 65–69% of the total variance in each season; pairwise distances in the projection approximately preserve those between the original error time series. Component model structures are grouped into mechanistic, statistical/ML, and hybrid approaches, shown in different colors. The official FluSight ensemble is shown using teal diamonds. Coral stars represent the embedding of perfect forecasts as a reference.

ILI forecasts also exhibited shared residual patterns, but were weaker and less stable (*Supporting Information, Extended structural-error analysis*, Fig. S1). This difference is consistent with ILI being a syndromic surveillance target rather than a pathogen-specific signal. The ILI activity reflects a mixture of influenza and other respiratory infections, as well as variation in healthcare-seeking behavior, outpatient reporting, and regional surveillance practices [18, 35, 36]. Thus, while ILI residuals contain some learnable short-horizon structure, they appear less coherent than hospitalization residuals, providing a useful contrast to assess when residual learning is most effective.

To quantify the diversity of component forecasts, we projected 0-week-ahead forecast error time series for hospitalizations into a two-dimensional representation using Principal Coordinates Analysis (PCoA, classical metric multidimensional scaling) [37] on pairwise Euclidean distances between the per-model error series (*Supporting Information, Principal coordinate analysis of forecast-error geometry*). PCoA is a metric dimensionality-reduction method that approximately preserves the original pairwise distances in the low-dimensional projection. As a reference, we also included the time series for perfect predictions, defined as zero forecast error at all time points.

Across all three seasons, the first two principal coordinates jointly explained approximately 65– 69% of the total variance in pairwise distances among models. Component models with different structures were embedded in relatively compact regions and mixed with one another, suggesting that forecast-error structure was not strongly determined by model class after calibration to observed data (Fig. 1D-F). The official FluSight ensemble was also located within the same region as the individual component models. In contrast, the embedding of perfect forecasts occupies a distinct position relative to the model-error cloud, indicating that the observed component-model errors are not randomly distributed around zero error. Together with the phase-dependent forecast error patterns in Fig. 1A-C, those patterns provided descriptive evidence of shared forecast-error structure and motivated the development of a residual-learning correction framework. See additional analyses for ILI and 2-week-ahead error structures in Figs. S2–S3.

### Seasonal-Temporal-Spatial Residual Learning framework

To correct the systematic errors identified above, we developed a Seasonal-Temporal-Spatial Residual Learning framework (STS-Residual), a post-prediction correction that learns structured forecast error from historical ensemble predictions. As shown in Fig. 2, STS-Residual learns the residual errors from three different components. (1) The seasonal component captures recurring epiweek-specific bias by fitting a statistical model to historical error patterns as a function of epidemic week, with recent-error regularization to adapt to current-season deviations. (2) The temporal component captures recent local error dynamics using a long short-term memory (LSTM) network [38], enabling the model to learn short-term dependencies in past forecasting errors in the same season. (3) The spatial component captures cross-location dependence in forecast error using a Graph Attention Network (GAT) [39] defined on a graph informed by location contiguity. These three components were then combined using an adaptive weighting mechanism that determines their relative contributions in each forecasting context.

**Figure 2:**
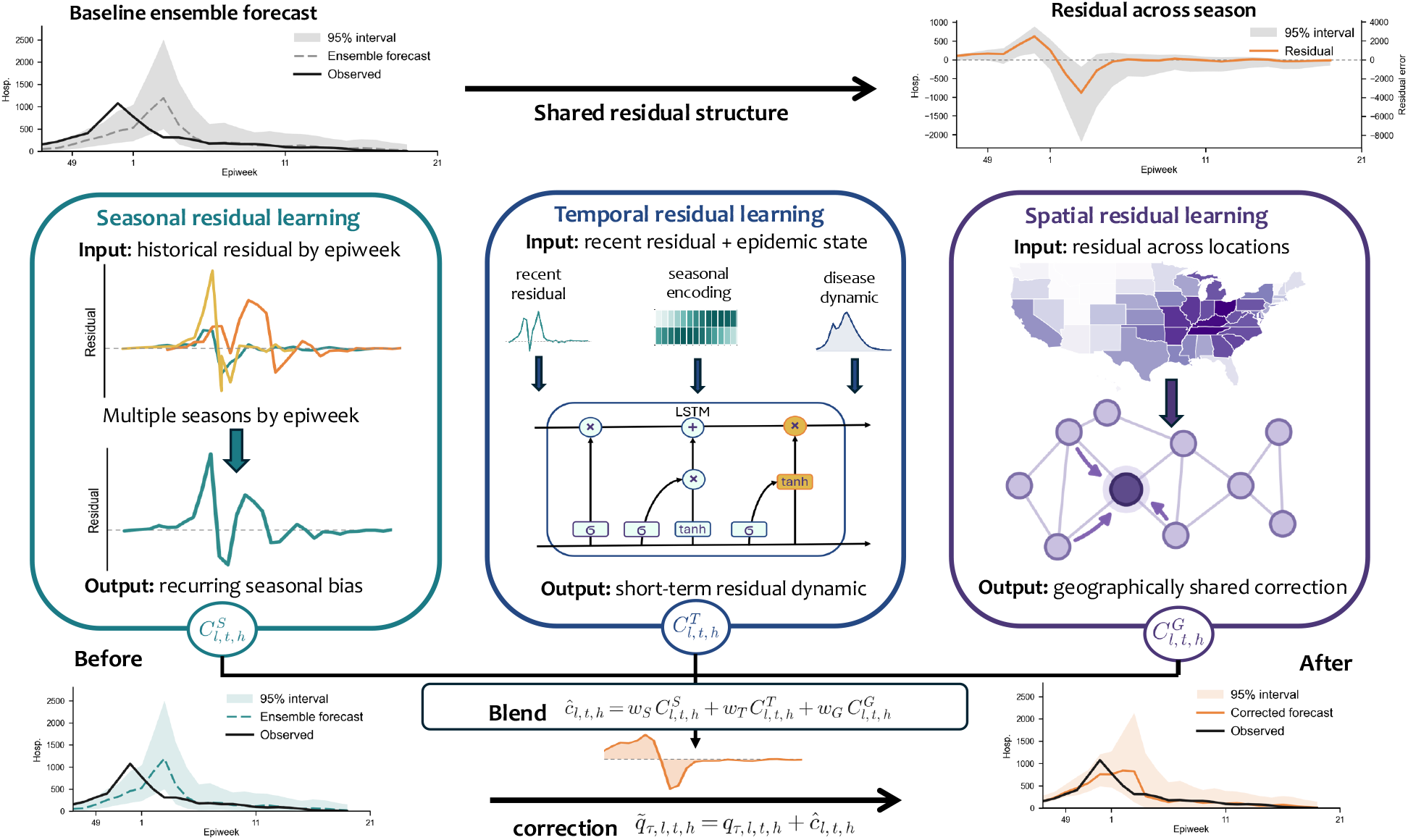
Conceptual overview of the Seasonal-Temporal-Spatial Residual Learning framework (STS-Residual). Historical ensemble forecasts are evaluated with observed outcomes to estimate residual errors, which exhibit shared seasonal, temporal, and spatial structure. STS-Residual models these systematic forecast errors through three components: 1) a seasonal learner that captures recurring epiweek-specific bias from historical residuals, 2) a temporal learner that models short-term residual dynamics using an LSTM, and 3) a spatial learner that captures cross-location residual dependence using a graph attention network (GAT). The three learned correction signals are adaptively integrated into a unified residual estimate, which is then added to the baseline probabilistic forecast.

For both forecast targets, STS-Residual was trained to learn residual errors from a training ensemble. For hospitalization forecasts, the training ensemble included the 12 retained models and their unweighted, quantile-wise mean; for ILI forecasts, it included all individual models and the constant-weight (CW) ensemble. The learned residual correction was then applied to the corresponding operational ensemble for evaluation. Specifically, for hospitalization, the correction learned from the retained-model training ensemble was applied to the official CDC FluSight ensemble, which could include additional eligible models not used to train STS-Residual. For ILI, the correction was applied to the CW ensemble distributed in the FluSight Network’s `cdc-flusight-ensemble` archive. In each case, the corrected forecast was compared with the same uncorrected operational ensemble used as the baseline. The correction was implemented by shifting every quantile of the operational probabilistic forecast by the same amount, thereby preserving the shape and width of the predictive distribution. Detailed model specifications and implementation are provided in Materials and Methods and *Supporting Information* (Tables S3–S4).

### The STS-Residual improves short-term forecasts

We applied the STS-Residual framework to hospitalization and ILI forecast data using leave-one-season-out (LOSO) cross-validation. For each testing season, STS-Residual was trained and validated on forecasts from all other seasons, with separate models fitted for each forecast horizon. To mimic real-time deployment, all residual inputs were restricted to lagged forecast errors for which the target observations would have been available before the correction was made. At each fore-casting week, we used the errors of the selected component models and the retained-model mean (training) ensemble up to that week as input to the trained STS-Residual model, which then generated a point prediction of error. This predicted error was subtracted from the official ensemble issued in the same week to produce a corrected prediction.

We used WIS and mean absolute error (MAE) to evaluate probabilistic and point forecast performance, respectively. Applying STS-Residual consistently reduced WIS for hospitalization forecasts across all four horizons relative to the official FluSight ensemble (Table 1). At the national level, WIS decreased by 19.9%, 12.5%, 9.9%, and 4.8% for horizons *h*=0 through *h*=3 weeks, respectively. These improvements were statistically significant for horizons *h*=0 through *h*=2 (*p <* 0.05, one-sided Wilcoxon signed-rank test). Pooled across states, WIS decreased by 11.6%, 6.9%, 5.0%, and 2.7% across the same horizons, with improvements significant at all four horizons (*p <* 0.001). MAE of the ensemble median showed a similar pattern. At the national level, MAE decreased by 22.3%, 13.1%, 8.6%, and 5.3% for horizons *h*=0 through *h*=3, respectively, with statistically significant improvements at *h*=0 through *h*=2 (*p <* 0.05). Pooled across states, MAE decreased by 10.3%, 6.0%, 3.9%, and 2.0%, with significant improvements at all four horizons (*p <* 0.001). Forecast calibration remained comparable: across sub-national locations, the corrected hospitalization forecasts achieved empirical coverage of 81–91% for the 95% prediction interval, compared with 81–90% for the FluSight ensemble; at the national level, coverage of the corrected forecast ranged from 77% to 96% across horizons.

**Table 1:** Forecast performance of STS-Residual. Forecasts corrected with STS-Residual (STS) are compared with the corresponding official benchmark ensemble (Bench.): the FluSight ensemble for hospitalization (Hosp.) and the constant-weight ensemble for ILI. Results are shown separately by forecast horizon. WIS evaluates probabilistic forecast accuracy, and MAE evaluates the accuracy of the median point forecast. Lower values indicate better performance. Improvement (Imp.%) is calculated as 100 *×* (1 − *M*_*ST S*_*/M*_*bench*_), where *M*_*ST S*_ and *M*_*bench*_ are the total WIS and MAE for the corrected and benchmark forecasts summed over all locations. Positive values indicate that STS-Residual performed better than the benchmark. Statistical significance was assessed with a one-sided Wilcoxon signed-rank test (sub-national: paired per-location mean scores; national: paired per-week scores). Coverage is the empirical proportion of observations contained within the 95% prediction interval. Pooled sub-national results include all reporting locations (*n* = 52 for hospitalization and *n* = 10 for ILI), and National refers to the U.S. national aggregate only. Hospitalization values are weekly admissions; ILI values are percentage points.

| Target | Scope | h | WIS |  |  |  |  | MAE |  |  |  |  | Cov 95% |  |
| --- | --- | --- | --- | --- | --- | --- | --- | --- | --- | --- | --- | --- | --- | --- |
| | | | Bench. | STS | $\Delta$ | Imp.% | p-value | Bench. | STS | $\Delta$ | Imp.% | p-value | Bench. | STS |
| Hosp. | Sub-national (52) | 0 | 44.4 | 39.2 | -5.2 | 11.6 | <0.001 | 65.8 | 59.0 | -6.7 | 10.3 | <0.001 | 90.2 | 90.7 |
|  |  | 1 | 70.8 | 65.9 | -4.9 | 6.9 | <0.001 | 103.8 | 97.6 | -6.2 | 6.0 | <0.001 | 86.5 | 85.3 |
|  |  | 2 | 95.5 | 90.8 | -4.8 | 5.0 | <0.001 | 138.0 | 132.6 | -5.3 | 3.9 | <0.001 | 84.3 | 83.4 |
|  |  | 3 | 117.8 | 114.6 | -3.2 | 2.7 | <0.001 | 167.0 | 163.7 | -3.3 | 2.0 | <0.001 | 81.4 | 80.7 |
|  | National | 0 | 1748.4 | 1400.7 | -347.7 | 19.9 | 0.002 | 2580.2 | 2004.5 | -575.7 | 22.3 | 0.001 | 88.4 | 95.7 |
|  |  | 1 | 2880.5 | 2519.2 | -361.3 | 12.5 | 0.017 | 4192.4 | 3644.1 | -548.3 | 13.1 | 0.010 | 83.6 | 87.3 |
|  |  | 2 | 4096.3 | 3689.3 | -407.0 | 9.9 | 0.038 | 5878.9 | 5373.9 | -505.0 | 8.6 | 0.046 | 81.3 | 83.4 |
|  |  | 3 | 5307.9 | 5055.6 | -252.3 | 4.8 | 0.130 | 7588.1 | 7187.5 | -400.6 | 5.3 | 0.084 | 78.7 | 77.4 |
| ILI | Sub-national (10) | 0 | 0.207 | 0.194 | -0.013 | 6.1 | <0.001 | 0.314 | 0.290 | -0.025 | 7.9 | <0.001 | 99.4 | 99.7 |
|  |  | 1 | 0.278 | 0.267 | -0.011 | 3.9 | 0.010 | 0.414 | 0.396 | -0.018 | 4.4 | 0.014 | 98.6 | 99.3 |
|  |  | 2 | 0.308 | 0.304 | -0.004 | 1.3 | 0.312 | 0.442 | 0.435 | -0.007 | 1.6 | 0.246 | 99.2 | 99.3 |
|  |  | 3 | 0.330 | 0.333 | +0.003 | -0.9 | 0.722 | 0.457 | 0.461 | +0.004 | -0.9 | 0.784 | 99.4 | 99.0 |
|  | National | 0 | 0.142 | 0.132 | -0.010 | 6.9 | 0.005 | 0.206 | 0.188 | -0.018 | 8.5 | 0.009 | 100 | 100 |
|  |  | 1 | 0.201 | 0.190 | -0.011 | 5.5 | 0.002 | 0.312 | 0.279 | -0.034 | 10.8 | 0.002 | 100 | 100 |
|  |  | 2 | 0.221 | 0.219 | -0.002 | 0.9 | 0.156 | 0.321 | 0.311 | -0.011 | 3.3 | 0.171 | 100 | 100 |
|  |  | 3 | 0.230 | 0.236 | +0.005 | -2.3 | 0.411 | 0.322 | 0.335 | +0.013 | -3.9 | 0.537 | 100 | 100 |

For ILI forecasts, improvements were more modest and concentrated at shorter horizons. Pooled across the 10 HHS regions, WIS decreased by 6.1%, 3.9%, and 1.3% for horizons *h*=0 through *h*=2, respectively, and was approximately unchanged at *h*=3 (−0.9%). These improvements were statistically significant only at the nowcast and one-week-ahead horizons (*p <* 0.05). MAE reductions showed a similar decline, from 7.9% at the nowcast horizon to near zero at longer horizons (Table 1; *Supporting Information*, Fig. S4). At the national level, improvements were likewise statistically significant at *h*=0 and *h*=1 (*p <* 0.05). This weaker performance is consistent with the less pronounced residual structure observed in ILI forecasts (*Supporting Information*, Fig. S1). Sensitivity analyses further showed that STS-Residual continued to improve forecasts when applied to an equal-weight hospitalization ensemble constructed from the retained models (*Supporting Information, Correction relative to the equal-weight mean ensemble*, Table S5), and that adding observed epidemic-state features yielded similar overall performance (*Supporting Information, Observed epidemic-state input features*, Table S6).

### Improvement across geographical locations

Measured by WIS, STS-Residual outperformed the FluSight ensemble in 48, 50, 51, and 48 of the 53 locations (national and sub-national) for horizons *h*=0 through *h*=3, respectively. The locations with poorer performance were generally low-incidence or geographically isolated, most notably Hawaii, Alaska, and Puerto Rico, where limited training signal may constrain the ability to learn stable residual patterns (*Supporting Information*, Table S7). The largest average reductions in MAE, approximately 8–12%, were observed in states including Missouri, Texas, Indiana, Michigan, Iowa, and Idaho (Fig. 3A–E). For ILI forecasts at the HHS-region level, geographic improvements were more limited and declined more strongly with forecast horizon: all 11 locations improved in WIS at the nowcast horizon, but only 4 of 11 remained improved by *h*=3 (Fig. 3F–J; *Supporting Information*, Table S8).

**Figure 3:**
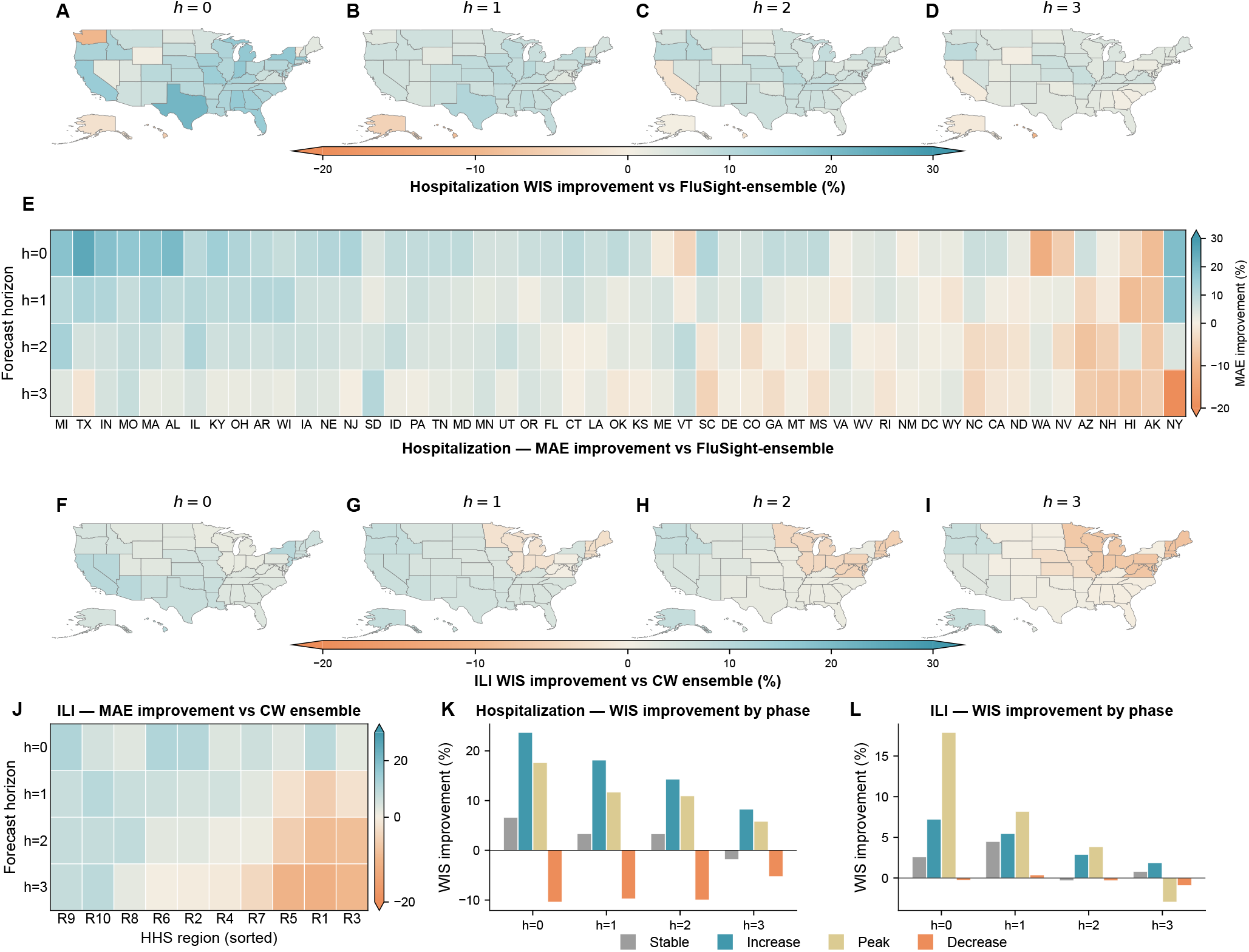
Performance of ensemble correction at sub-national scales and across epidemic phases. (A-D) WIS improvement for influenza hospitalization forecasts relative to the FluSight ensemble across U.S. states and jurisdictions at four forecast horizons. Colors indicate the percentage reduction in WIS, with teal denoting improved performance. Percentage changes in WIS were averaged across three seasons. (E) MAE reduction for influenza hospitalization forecasts. Each column represents one state, and rows indicate forecast horizons. States are ordered from left to right by decreasing average MAE reduction. (F-I) WIS improvement for ILI forecasts relative to the FluSight constant-weight (CW) ensemble across U.S. HHS regions at four forecast horizons. Colors indicate the percentage reduction in WIS, with teal denoting improved performance. Percentage changes in WIS were averaged across seven seasons. (J) Percentage reduction in MAE for ILI forecasts across the 10 HHS regions, averaged over seven seasons. Regions are ordered from left to right by decreasing improvement. (K) Percentage reduction in WIS for hospitalization forecasts across epidemic phases, averaged over all states and seasons. (L) Percentage reduction in WIS for ILI forecasts across epidemic phases.

### Stronger improvement during epidemic growth and around peaks

For each location and season, we stratified epiweeks by epidemic phase. *Peak* weeks were defined as weeks in which the observed value reached at least 85% of the season- and location-specific maximum. The remaining weeks were classified as *Increase* or *Decrease* if the week-over-week change exceeded 5% of the current level, and as *Stable* otherwise. For hospitalizations, WIS decreased by 8-24% during the *Increase* phase and 6-18% during *Peak* weeks, largest at the nowcast horizon and diminishing at longer leads, whereas *Stable* weeks showed only small changes (up to 7%); during the *Decrease* phase the correction tended to overcorrect, increasing WIS by 5-10% (Fig. 3K). For ILI, gains were concentrated at the nowcast horizon, reaching 18% during *Peak* weeks and 2-7% during *Increase*, and were near zero during the *Stable* and *Decrease* phases (Fig. 3L).

### STS-Residual component ablation analysis

We further performed an ablation analysis to quantify the contribution of each STS-Residual branch (*Supporting Information*, Table S9). For hospitalization forecasts, the seasonal branch accounted for most of the improvement, especially at the nowcast horizon, while the temporal and spatial branches provided modest additional gains at longer horizons; the full adaptive blend gave the best mean MAE improvement, though its margin over the seasonal branch alone was small. In contrast, for ILI forecasts, most of the improvement was explained by the seasonal branch: seasonal-only and seasonal-containing configurations performed similarly to the full model, whereas the temporal and spatial branches provided limited additional signal. These results show that the adaptive blend can route weight toward the most informative residual components in each forecasting context, allowing the same framework to adapt to datasets with different spatial resolutions and error structures.

### Robustness to changing component-model composition

Real-time forecast hubs experience shifting team participation across seasons, so the operational ensemble in any given week is built from the models available at that time. To test whether the gains from STS-Residual depend on a fixed pool of component models, we performed a Monte Carlo transfer experiment. At each of 10 iterations, we drew an *independent* random subset of 7 of the 9 individual-team hospitalization models *separately for every season*, rebuilt that season’s equal-weight ensemble from its own subset, and trained STS-Residual under the same LOSO protocol. The corrector trained on the training-season subsets was then applied to the held-out season’s subset ensemble, whose composition differs from those used in training, and evaluated against that same uncorrected subset ensemble. To make the corrector agnostic to model identity, the temporal and spatial branches used only ensemble-level inputs (the ensemble residual and the cyclic epiweek encoding). Because the rebuilt subset ensembles were occasionally unstable, each branch’s correction was passed through an overcorrection safeguard that capped its normalized magnitude (Materials and Methods).

Across the 3 *×* 10 = 30 season-level validation runs, STS-Residual improved upon the subset-rebuilt ensemble at every horizon, with mean WIS reductions of 12.1%, 7.6%, 7.3%, and 4.6% for horizons *h*=0 through *h*=3, respectively. MAE reductions were 14.6%, 7.0%, 7.1%, and 4.6%, with corrected forecasts outperforming the uncorrected baseline in 90-100% of runs at each horizon (*Supporting Information, Robustness to changing component-model composition*, Table S10). This consistently positive transfer to ensembles built from model sets not seen during training indicates that the learned residual structure is shared across ensemble compositions rather than tied to specific component-model identities.

### Robustness to real-time surveillance data backfill

In real-time surveillance, initially reported data are often updated in subsequent releases [40], a process known as backfill. Real-time forecasting systems must therefore generate predictions using the initially reported data available at the forecast date. To assess whether the gains from STS-Residual are robust to these real-time data revisions, we used the two hospitalization seasons, 2024-25 and 2025-26, for which the NHSN data provide both first-released and finalized weekly hospitalizations. We re-ran the full pipeline using the LOSO design, computing lagged residual features in the held-out season relative to the *first-released* observations while retaining finalized observations for the training seasons. The same overcorrection safeguard was applied. We evaluated the corrected forecasts under two scoring conventions: against the first-released targets, representing the surveillance values available in real time, and against the finalized targets, representing the eventually revised observations.

Pooled across both seasons and all 53 locations, the corrected ensemble reduced WIS relative to the FluSight ensemble at every horizon under both conventions (Table S11). WIS reductions were 12.6%, 9.6%, 4.7%, and 1.6% for horizons *h*=0 through *h*=3 when scored against first-released targets, and 13.1%, 9.2%, 4.7%, and 1.7% when scored against finalized targets. MAE reductions are reported in the *Supporting Information, Robustness to real-time surveillance revisions*. Although first-released and finalized admissions differed in 74% of scored location-weeks, the estimated WIS improvements differed by no more than 0.5 percentage points between the two scoring conventions. These results indicate that the benefit of STS-Residual is not particularly sensitive to backfill.

## Discussion

Model diversity is central to the success of ensemble forecasting. A common expectation is that combining models with different structures, data streams, and calibration strategies can reduce idiosyncratic errors and improve predictive robustness. Our results show, however, that component models can share systematic forecast errors despite substantial methodological diversity. These shared errors were especially pronounced for influenza hospitalization forecasts, where models tended to underestimate during epidemic growth and overestimate after seasonal peaks. Such patterns likely arise, at least in part, from a common challenge faced by all real-time forecasting approaches: models must infer changing epidemic dynamics from surveillance data that are incomplete, noisy, and dominated by past observations. During periods of rapid epidemic growth or near turning points, the most informative observations about changing transmission dynamics represent only a small fraction of the available training data. As a result, models may tend to smooth over or lag behind emerging changes, even when they differ in structure or fitting procedure. This mechanism is consistent with principles from data assimilation [41], in which updated estimates balance prior model states against new observations according to their respective uncertainties. Such structural lag cannot be fully eliminated by adding more component models or by reweighting an ensemble. Our findings, therefore, highlight a fundamental limitation of ensemble epidemic forecasting: forecast diversity, rather than simply the number of models, is critical. At the same time, we show that these shared errors are not random. When residual patterns are sufficiently strong and recurrent, they can be learned from past forecasts and used to improve real-time predictions.

Residual learning provides a natural framework for addressing this challenge. Rather than replacing existing forecasting models, residual learning treats their remaining errors as an additional modeling target. This idea has been widely used in machine learning [42, 43], where models often learn corrections to an initial prediction. Similar ideas have also been applied in epidemic forecasting. For instance, previous work has learned model residuals for a mechanistic epidemic model to improve forecasting [44]. Our approach differs by learning residual structure shared across an ensemble of component models, rather than errors tied to a single forecasting system. This distinction is important for operational forecasting, where the set of participating models can change across seasons and weeks. Our component-model composition experiments further suggest that ensemble-level residual patterns can remain informative when the set of contributing forecasts changes.

The contrast between hospitalization and ILI forecasts helps clarify when residual learning is most useful. Hospitalization is a more specific and severe outcome, with larger variation in magnitude across locations and epidemic phases than ILI. This larger dynamic range may make rapid changes harder for component models to track in real time, strengthening the shared lagged error structure during growth and peak periods. In contrast, ILI is a syndromic target that combines influenza with other respiratory infections and is affected by healthcare-seeking behavior, outpatient reporting, and surveillance practices. These factors likely weaken and destabilize shared residual patterns, consistent with the more modest improvements observed for ILI, especially at longer horizons. Thus, residual learning is most valuable when ensemble errors are coherent, stable, and operationally observable, as in influenza hospitalization forecasting.

Several limitations should be noted. First, our evaluation used archived real-time forecasts. Although this design closely mimics prospective forecasting, the framework should be further tested in fully operational real-time settings. Second, the available hospitalization forecast record covered only three seasons, limiting the range of epidemic dynamics available for training and validation. The strong improvements in hospitalization are encouraging, but additional seasons will be important for assessing robustness to unusual epidemic timing, changing reporting practices, and shifts in model participation. Third, the performance of STS-Residual depends on the quality of the underlying component forecasts and on the availability of sufficient historical data. In settings with sparse surveillance, limited model participation, or highly irregular dynamics, residual structure may be harder to learn. Fourth, residual correction introduces the possibility of overcorrection, particularly immediately after turning points or when the current season deviates from historical patterns; this was reflected in poorer performance during some decreasing-phase weeks. Finally, this framework may be most effective for pathogens with recurring seasonal structure, such as influenza in temperate regions. Its performance may be reduced for diseases or regions with less regular epidemic dynamics, such as influenza in tropical settings. Future work should evaluate adaptive safeguards against overcorrection and extend residual learning to broader disease systems and surveillance contexts.

## Materials and Methods

### Forecast data

We analyzed two retrospective FluSight forecasting datasets that differ in target outcome, spatial resolution and historical coverage. The first consisted of probabilistic forecasts of laboratory-confirmed weekly influenza hospital admissions for the 2023–24, 2024–25 and 2025–26 seasons. Forecasts and subsequently observed outcomes were obtained from the CDC FluSight forecast hub [2] and `HealthData.gov`. We retained the 12 models that submitted forecasts for all three seasons at horizons *h* ∈ {0, 1, 2, 3} across 53 locations (50 states, the District of Columbia, Puerto Rico, and the U.S. national aggregate), including models submitted by individual teams and the CDC ensembles. The list of models appears in *Supporting Information*, Table S1. The probabilistic forecasts for short-term hospitalizations include 23 quantiles of the predictive distribution {0.010, 0.025, 0.050, 0.100, 0.150, · · ·, 0.950, 0.975, and 0.990}.

The second dataset consisted of influenza-like illness forecasts from the 2010–11 through 2016–17 seasons, together with the corresponding weighted influenza-like illness observations. These forecasts were obtained from the FluSight Network `cdc-flusight-ensemble` archive [3, 5] and covered the ten Department of Health and Human Services regions, with forecasts issued weekly from epiweek 40 through epiweek 20. Because ILI component models report predictive distributions as categorical probabilities over fixed 0.1-percentage-point bins rather than as 23 quantile levels, we converted bin probabilities to 23 FluSight quantiles by linear interpolation of the cumulative distribution, discarding forecasts whose total probability fell outside [0.9, 1.1] and renormalizing the remaining bins to unit mass; forecasts were further excluded for any (season, location, horizon) combinations with fewer than five valid weekly submissions.

### Training ensembles and residual targets

For each forecast target, we first constructed a training ensemble from the retained model submissions and used its residuals as the learning target for STS-Residual. Let *m* ∈ {1, …, *M}* index the retained component models, let ens denote the training ensemble associated with those models, and let 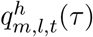 denote the *τ*-quantile forecast from the component model *m* for location *l*, forecast issue week *t*, and horizon *h*, where the target is the observed value *y*_*l,t*+*h*_. The same notation is used for the training ensemble forecast, 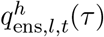. For hospitalizations, the training ensemble was constructed as the equal-weight, quantile-wise mean of the 12 retained models described above. For ILI, we directly used the constant-weight (CW) ensemble distributed with the FluSight Network’s `cdc-flusight-ensemble` archive [3, 5] as the training ensemble. The CW ensemble combines the 21 component models with weights that are held constant across the weeks of each season but estimated separately per season, so a few models dominate while many receive near-zero weight. We adopted it because it attained the best retrospective skill among the archive’s pre-computed ensembles. The ensemble median 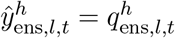 (0.5) was used to define the residual learning target. Residuals are normalized by a fixed, exogenous location scale *S*_*l*_ before learning and rescaled afterward, so that the learners operate on a comparable scale across jurisdictions whose target magnitude differs by orders of magnitude:

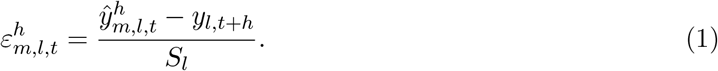

The same definition applies to the training ensemble (set *m* = ens), giving the ensemble residual 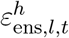 that STS-Residual learns to predict and correct. Positive residuals, therefore, indicate overprediction and negative residuals indicate underprediction. For hospitalizations, *S*_*l*_ was defined using the location population (in 100,000 persons), yielding a per-capita residual scale. For ILI, where the outcome is already expressed as a population-weighted percentage, we set *S*_*l*_ = 1.

### Seasonal–Temporal–Spatial Residual Learning framework

The objective of STS-Residual is to learn a correction 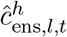 that approximates the negative of the expected residual error and then add this correction to the original ensemble forecast. To achieve this, STS-Residual decomposes forecast error into three complementary sources: 1) recurrent seasonal residuals, 2) temporal dynamic residuals, and 3) spatially dependent residuals. This design directly follows the empirical observation in Fig. 1 that component models with different structures nevertheless share similar error trajectories, particularly during epidemic growth and seasonal peaks.

#### Seasonal residual learning

The seasonal component captures systematic residuals associated with influenza season timing. On each training fold we estimate the mean normalized training ensemble (ens) residual for each location *l* at epiweek *e* and horizon *h*:

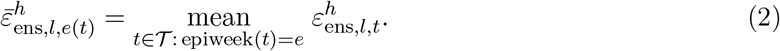

Here, *T* denotes the set of location–week observations for seasons in the training data and *e*(*t*) is the epiweek of week *t*. The estimated value 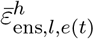 was then assigned to any forecast issued in the same epiweek *e*, yielding a historical estimate of recurrent epiweek bias. Because epidemic dynamics vary across seasons, relying only on these historical epiweek patterns could over-correct when the current season is shifted earlier or later. We therefore regularized the seasonal residual using a recent local error, defined as the average of the most recent horizon-0 residuals in prior weeks:

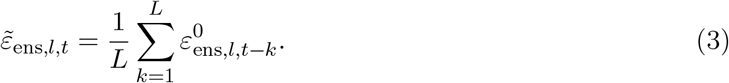

Here, *L* is the number of recent weeks to consider. The estimated value 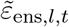 provides a local real-time adjustment for deviations of the current season from the historical epiweek pattern. The seasonal correction is the convex combination of these two residuals:

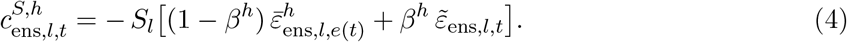

The negative sign ensures that the correction offsets the estimated residual when added to the forecast. The mixing weight *β*^*h*^ ∈ [0, 1] and *L* ∈ {1, …, 8} are selected per horizon by inner cross-validation on the training fold.

#### Temporal residual learning

The temporal component captures local short-term error dynamics that may not be fully explained by the seasonal residual alone. The temporal component models the short-term dynamics of the residual with a long short-term memory network (LSTM) [38]. To mimic real-time forecasting, the temporal learner uses only residuals from forecasts whose target observations would have been available at the time the correction is made.

At each reference week *t*, location *l*, and forecast horizon *h*, we construct an input sequence of length *W* whose timestep *i* ∈ {1, …, *W}* carries the feature vector. Because a separate LSTM is trained per horizon, the temporal learner reads a length-*W* window of the most recent *nowcast* (*h*=0) residuals, shared across all target horizons. The *h*=0 residual 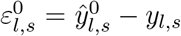 depends only on the observation at week *s*, so it is available whenever *s* ≤ *t* − 1 and the window is leak-free for every horizon. Let *s*_*i*_ = *t* − *W* − 1 + *i, i* = 1, …, *W* index the *W* most recent issue weeks before *t* (so *s*_1_ = *t* − *W*, …, *s*_*W*_ = *t* − 1). The feature vector at time step *i* is

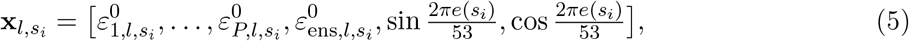

The leading *P* entries are the nowcast residuals of the individual input models, followed by the now-cast residual of the training ensemble and a cyclic encoding of seasonal position; the same window maps to a horizon-specific correction because a separate LSTM is fit for each *h*. For hospitalizations, the input models are the retained component models excluding the FluSight-ensemble (which is reserved as the evaluation benchmark and is therefore not used as an input feature), giving *P* = 11; for ILI, all retained component models are used, giving *P* = 21.

The sequence 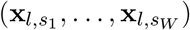 is fed to an LSTM and the terminal hidden state is mapped through a multilayer perceptron to produce a scalar estimate of the normalized training ensemble residual. The temporal correction is then given by:

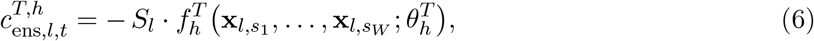

where 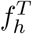 denotes the LSTM-based temporal residual learner. Implementation details, including the LSTM architecture, training procedure, optimization settings, and horizon-specific hyperparameter selection, are provided in *Supporting Information, STS-Residual implementation details*.

### Spatial residual learning

The spatial component captures the fact that forecast errors may be shared across geographically connected locations because neighboring jurisdictions often experience related epidemic dynamics, surveillance artifacts, and timing shifts. We model this structure with a graph attention network (GAT) [39], allowing each location to borrow information from geographically contiguous neighbors through learned, context-specific attention weights. We also evaluated a graph structure informed by mobility between locations using flight and cellphone-based foot-traffic data [45], but this alternative yielded lower performance.

We define an undirected graph *G* = (*V, E*): for hospitalizations *V* comprises the 50 states, the District of Columbia, and Puerto Rico (52 nodes); for ILI, it comprises the ten HHS regions and the U.S. national aggregate (11 nodes). For hospitalizations, edges connect locations that share a land border; Alaska, Hawaii, and Puerto Rico therefore have only self-loops. For ILI, edges connect HHS regions that share a border. Every node carries a self-loop so that each location can retain its own residual history. Unlike the temporal branch, a *single* GAT is trained jointly across all four horizons (full-graph snapshots are scarce, ≈55 per fold, so a per-horizon model is infeasible); it therefore uses the *horizon-specific* lagged residuals *ε*^*h*^ rather than the nowcast residuals, which is what lets one shared model distinguish the four lead times. For target horizon *h* these are taken from the shifted window 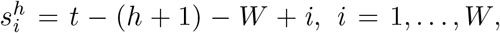, whose most recent target falls at week *t* − 1 and is thus leak-free. At each reference week *t* and horizon *h*, this residual sequence for each location is first encoded by a location-shared LSTM, yielding a node-level representation 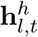. These representations are passed to a multi-head GAT layer, which adaptively aggregates information from each node’s neighboring locations *N*(*l*). The spatial module then maps the aggregated neighborhood representation to a scalar estimate of the normalized training ensemble residual. The corresponding spatial correction is

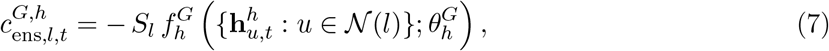

where 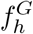 denotes the GAT-based spatial residual learner. Details of the attention mechanism, graph construction, and training procedure are provided in *Supporting Information, STS-Residual implementation details*.

### Adaptive blend

The three component corrections are combined by a per-horizon convex blend,

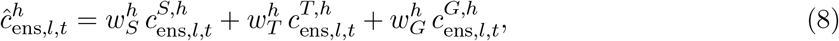

where 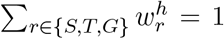 and 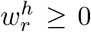. The weights were selected on a 0.05 grid by inner cross-validation on the training fold. Before blending, each branch’s normalized correction 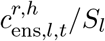 (*r* ∈ *{S, T, G}*) is clipped to [−1, 1] and rescaled by *S*_*l*_, an overcorrection safeguard that bounds each branch’s shift to one per-capita scale unit; this safeguard is applied throughout. The corrected forecast is obtained by shifting every quantile of the predictive distribution by 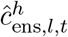, clipping at zero, and enforcing monotonicity; we impose no further parametric assumption on the residual distribution.

### Experimental setting and evaluation metrics

All evaluations use leave-one-season-out (LOSO) cross-validation: each fold treats one season as the test set, with the remaining seasons used for training and inner cross-validation, and the rotation is repeated until every season has served as the test exactly once. All hyperparameters were selected separately for each horizon by inner cross-validation within the training fold.

Corrected forecasts were evaluated against each dataset’s official ensemble. For hospitalizations, the primary benchmark was the CDC FluSight-ensemble, the officially published forecast formed each week as the quantile-wise median across eligible submissions; as a secondary comparison (*Supporting Information*) we benchmarked against the equal-weight mean of the 12 selected models. For ILI, the benchmark was the CDC constant-weight ensemble distributed with the FluSight Network archive [5].

The primary metric is the Weighted Interval Score (WIS) [46], evaluated on the 23 quantile levels *{*0.01, 0.025, 0.05, …, 0.95, 0.975, 0.99*}* after shifting the ensemble’s quantiles by the learned 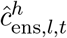. We also report the Mean Absolute Error (MAE) of the median forecast and empirical coverage at the 95% prediction intervals. Performance is summarized relative to each dataset’s CDC official ensemble (FluSight-ensemble for hospitalizations, constant-weight ensemble for ILI), with positive values indicating improvement after correction. In the main analysis the learned correction is applied to each target’s official ensemble; in the *Supporting Information, Correction relative to the equal-weight mean ensemble*, we additionally apply the same correction to the retained-model mean ensemble and report its improvement over that uncorrected ensemble.

Statistical significance of the WIS and MAE reductions was assessed for each target and horizon with a one-sided Wilcoxon signed-rank test (corrected vs. uncorrected). For the sub-national scope, the test used the paired per-location mean scores (*n* = 52 for hospitalization, *n* = 10 for ILI); for the national aggregate, which is a single series, the test used the paired per-week scores across all weeks and seasons. *p*-values are reported in Table 1.

### Robustness analyses

We performed two robustness analyses for the hospitalization target, both reusing the main training pipeline.

First, we evaluated robustness to changes in component-model composition. At each of 10 iterations, we drew an independent random subset of 7 of the 9 individual-team models separately for every season (training and held-out), rebuilt that season’s equal-weight ensemble from its own subset, and trained STS-Residual under the same leave-one-season-out protocol. The trained corrector was applied to the held-out season’s subset ensemble, whose composition differs from those in the training seasons, and compared against that same uncorrected subset ensemble. To make the corrector agnostic to model identity, the LSTM and GAT branches used only ensemble-level inputs (the ensemble residual and the cyclic epiweek encoding) rather than the per-model residuals used in the main analysis. Because subset-rebuilt ensembles are occasionally unstable, each branch’s normalized correction *c/S*_*l*_ was clipped to [−1, 1] before the adaptive blend, the same overcorrection safeguard used in the main analysis. Detailed settings and results are in the *Supporting Information*.

Second, we evaluated robustness to real-time surveillance revisions. Confirmed influenza admissions reported through NHSN are revised after first publication; for the two seasons with available revision history (2024–25 and 2025–26) we obtained the first-released and finalized weekly values from the Delphi Epidata API [47]. We re-ran the main pipeline (LSTM, GAT, and adaptive blend) holding out one season at a time: the held-out season’s residual features were computed against its *first-released* observations while the training seasons retained finalized values, and the corrected forecast was scored two ways, against the first-released and against the finalized targets. The same [−1, 1] overcorrection safeguard was applied, and improvement is reported pooled over both seasons and all 53 locations relative to the FluSight-ensemble. Detailed settings and results are in the *Supporting Information*.

## Data availability

All analysis and figure-generation code, together with the scripts used to fetch and process the third-party data, is available on GitHub (https://github.com/Jensen6667/sts-residual). This study uses publicly available third-party data that we do not redistribute: influenza hospitalization and influenza-like illness forecasts from the CDC FluSight forecast hub and the FluSight Network `cdc-flusight-ensemble` archive; laboratory-confirmed influenza hospital admissions from NHSN, obtained (including first-released and revised vintages) through the Delphi Epidata API [47]; and hospitalization ground-truth data from `HealthData.gov`. Scripts and instructions for retrieving each source are provided in the repository.

## Author contributions

S.P. and H.D. designed research; Y.Q. performed analysis; Y.Q., H.D., and S.P. investigated results; and Y.Q., H.D., and S.P. wrote the paper.

## Acknowledgments

This work was supported by NIH grants R35GM156799, R01AI63023, NSF grant DMS-2229605, and Columbia University Research Stabilization Fund.

## Supporting Information

### Ensemble errors for individual models with correlated errors

Let *e*_*i*_ denote the forecast error from component model *i*, for *i* = 1, …, *N*. The ensemble error is defined as the average error across component models:

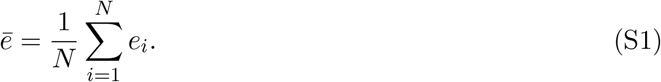

Assume that each component model has the same error variance,

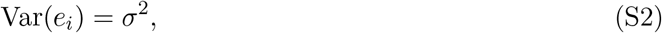

and that the average pairwise correlation between component model errors is *ρ*. Thus, for *i* ≠*j*,

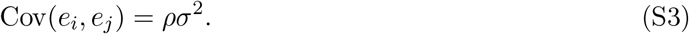

The variance of the ensemble error is then

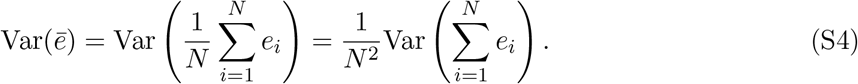

Using the variance-of-a-sum identity,

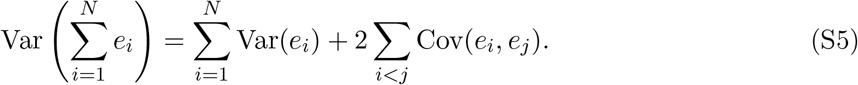

Because there are *N* variance terms and *N* (*N* − 1)*/*2 covariance terms,

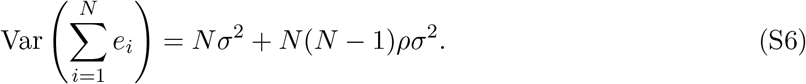

Therefore,

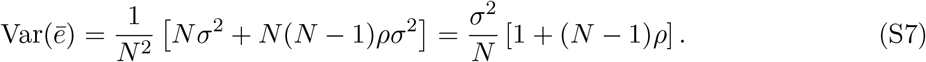

Equivalently,

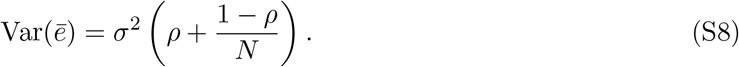

Thus, the standard deviation of the ensemble error is

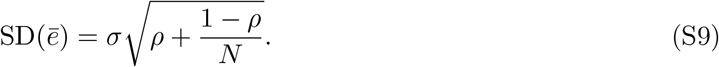

As *N* → ∞,

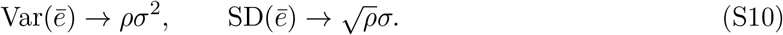

Therefore, when component model errors are positively correlated, adding more models reduces only the independent component of error, while the shared error component remains.

### Retained forecast models

For **hospitalizations**, we retain the 12 component models in the CDC FluSight hub [2] that submitted forecasts in every season from 2023–24 to 2025–26 across all four target horizons (Table S1). These included 9 individual team submissions and 3 hub-generated reference forecasts: *FluSight-ensemble* (quantile-wise median of designated submissions), *FluSight-lop_norm* (linear opinion pool of designated submissions), and *FluSight-baseline* (persistence of the most recent observation). The equal-weight, quantile-wise mean of all 12 retained forecast series was used as the hospitalization retained-model training ensemble whose residuals were learned by STS-Residual. *FluSight-ensemble* is used throughout the paper as the benchmark against which the STS-Residual correction is evaluated and is therefore excluded from the LSTM and GAT input features. The remaining 11 retained models—the 9 individual-team submissions, the *FluSight-baseline* persistence reference, and the *FluSight-lop_norm* aggregate—are used as the per-model input features. A more restrictive 9-model team-only pool (excluding both *FluSight-baseline* and *FluSight-lop_norm*) is used for the random-subset robustness analysis described in Section “Robustness to changing component-model composition”.

**Table S1:** Twelve models retained for the hospitalization analysis. All twelve submitted forecasts in every season (2023–24, 2024–25, 2025–26) for horizons *h* = 0, 1, 2, 3. Team affiliations and method summaries are taken from each model’s metadata file in the FluSight-forecast-hub repository.

| Model identifier | Team / institution | Approach (summary) | Type |
| --- | --- | --- | --- |
| CEPH-Rtrend_fluH | CEPH Lab, Indiana University | Renewal equation with Bayesian $R_t$ estimation | Mechanistic |
| CMU-TimeSeries | Delphi (CMU) | Quantile auto-regression ensemble | Statistical |
| CU-ensemble | Columbia University | Weighted ensemble of SIR-EAKF, drift, GBM, DL | Hybrid |
| FluSight-baseline | CDC FluSight hub | Persistence of most recent observation | Baseline |
| FluSight-ensemble | CDC FluSight hub aggregate | Quantile-wise median of designated submissions | Hub ensemble |
| FluSight-lop_norm | CDC FluSight hub aggregate | Linear opinion pool of designated submissions | Hub ensemble |
| MIGHTE-Nsemble | MIGHTE Lab | Adaptive WIS-weighted ARIMA + LightGBM + SVM | Statistical |
| PSI-PROF | Predictive Science Inc. | SIR[H]2 mechanistic model with MCMC inference | Mechanistic |
| SigSci-TSENS | Signature Science | Equal-weight time-series ensemble | Statistical |
| UM-DeepOutbreak | University of Michigan | Seq2seq RNN/attention with conformal prediction | Deep learning |
| UMass-flusion | Reich Lab (UMass) | Spatial-aware statistical + ML ensemble | Hybrid |
| UVAFluX-Ensemble | University of Virginia (Biocomplexity) | Multi-method statistical + DL ensemble | Hybrid |

For **influenza-like illness (ILI)**, the FluSight Network’s `cdc-flusight-ensemble` historical archive [3, 5] contains contributions from 21 individual component models across seasons 2010–11 through 2016–17 (Table S2). The constant-weight (CW) ensemble shipped with the archive is used as the training ensemble whose residuals were learned, the ensemble to which the correction was applied, and the benchmark for evaluating the corrected forecasts.

### Principal coordinate analysis of forecast-error geometry

To quantify the similarity among component models’ real-time forecast errors, we embed their per-model error time series with classical Principal Coordinates Analysis (PCoA, also known as classical metric or Torgerson multidimensional scaling) [37], which arranges the models in a low-dimensional space whose pairwise distances approximate the original ones.

For a given season and horizon *h*, each retained component model *m* contributes the time series of its US-national forecast errors 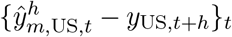 over that season’s forecast weeks (we use the nowcast horizon *h* = 0 in Fig. 1D–F and repeat the analysis at *h* = 2 in Figs. S2 and S3). A synthetic “Perfect” series of all zeros is added as a fixed zero-error reference, and weeks with no submission are set to zero error so that every series has the same length.

**Table S2:**
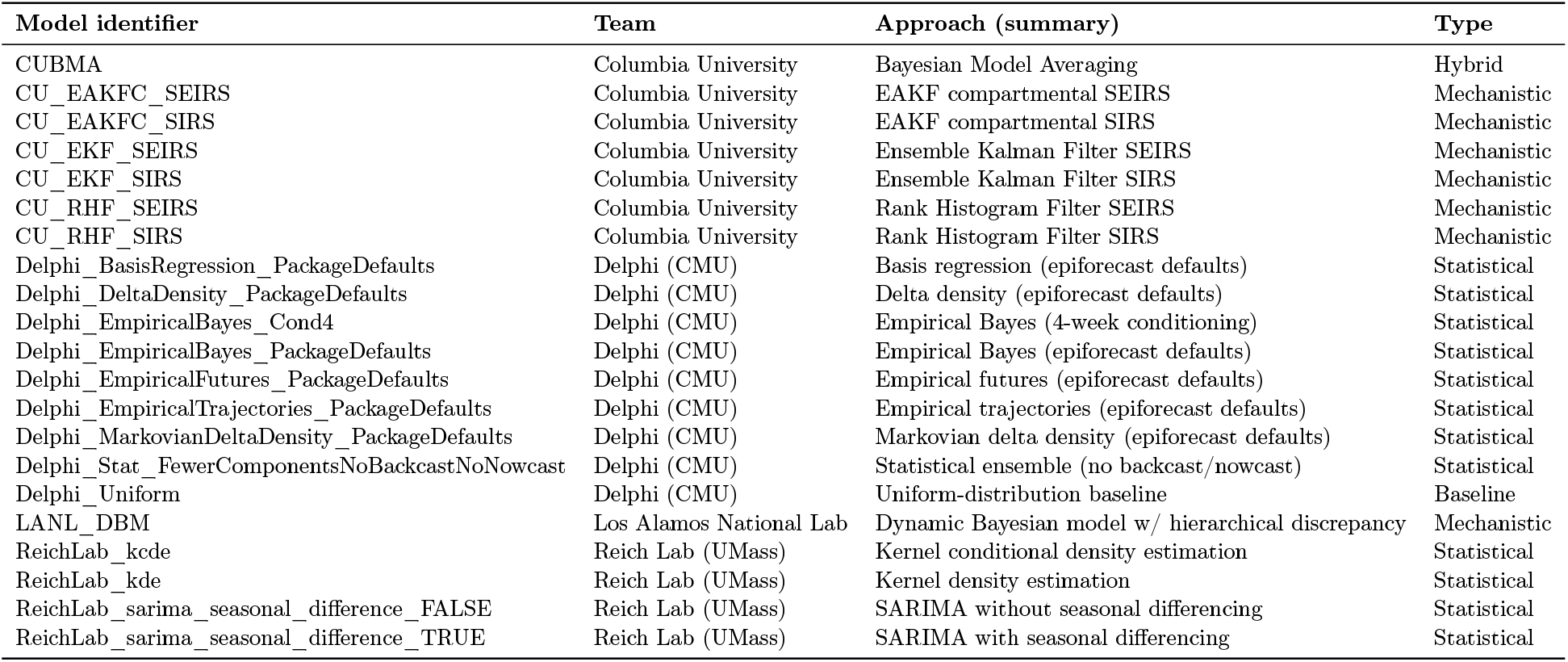
Twenty-one component models contributed to the FluSight Network cdc-flusight-ensemble archive across the seven ILI seasons (2010–11 through 2016–17). Affiliations and model names taken from the `metadata.txt` of each component-model folder in the archive.

Let *D* be the matrix of pairwise Euclidean distances among the forecast-error time series (the component models together with the Perfect reference). We obtain classical PCoA coordinates from *D* by first constructing the squared-distance matrix *D*^(2)^, with entries 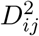. We then double-center this matrix to obtain:

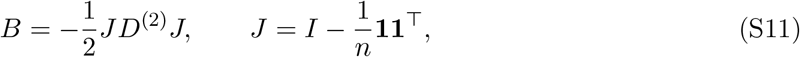

where *n* is the number of series. The matrix *B* is then eigendecomposed as

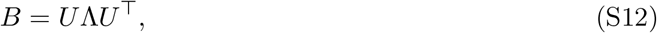

with eigenvalues ordered in decreasing magnitude. The first two principal coordinates are given by 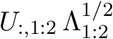. We report the fraction of positive PCoA inertia explained by these two axes,

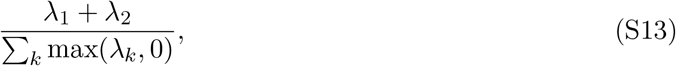

which ranges from 65–69% across the hospitalization seasons and is shown separately in each panel of Fig. 1. Because PCoA preserves the input distances, the spread among the model points and their separation from the Perfect reference are on the same scale as the original error series; that separation runs mainly along PCo1, the axis carrying the largest share of distance variance and— because the Perfect series has zero magnitude—the one most aligned with overall error size. Each season is analyzed independently, so within-season error structure is never conflated across seasons.

## Extended structural-error analysis

To confirm that the structural-error pattern motivating STS-Residual (Fig. 1 of the main text) is not specific to a single dataset or horizon, we provide three additional renderings of the component-model error matrix. Figure S1 reproduces Fig. 1 for the seven ILI seasons at the nowcast horizon (*h* = 0): the same tight clustering of all 21 component models, with the synthetic “Perfect” (zero-error) point clearly separated in the PCoA, is observed in every season. Figure S2 and Fig. S3 extend the analysis at the longer 2-week-ahead horizon (*h* = 2). At this longer horizon, the retained forecasts continued to exhibit overlapping error geometries, although the degree of clustering and separation from the zero-error reference varied across seasons and targets.

Taken together, the PCoA analyses provide descriptive evidence that forecasts produced by different modeling approaches can occupy similar regions of forecast-error space. For hospitalization, the component models and the official FluSight ensemble form a relatively compact cloud that is consistently displaced from the synthetic “Perfect” zero-error reference across seasons. The ensemble lies within the same model cloud rather than between the cloud and the zero-error point, indicating that aggregation of the submitted forecasts inherits a shared component of error. In this setting, re-weighting the existing component forecasts can reduce idiosyncratic disagreement among models, but it is unlikely to remove a systematic offset that is not represented by the model pool itself. This provides the clearest empirical motivation for STS-Residual: the correction must act in the residual direction between the ensemble forecast and the observed outcome, rather than simply redistributing weight among existing submissions.

The ILI forecasts exhibit a weaker and more heterogeneous version of this pattern. Across ILI seasons, the component models still tend to cluster in PCoA space, indicating correlated error trajectories, but the Perfect reference is sometimes located near or within the component-model cloud. This suggests that, for some ILI seasons, the zero-error direction is at least partially represented among the submitted models, so an appropriately weighted ensemble could, in principle, reduce error more effectively than in the hospitalization setting. Thus, the structural offset is stronger and more visually consistent for hospitalization, whereas ILI shows greater season-to-season variability and more opportunity for error cancellation through aggregation. STS-Residual is therefore motivated by the same principle in both datasets, but the expected gains are naturally larger when the existing model pool is collectively displaced from the truth, as observed for hospitalization.

**Figure S1:**
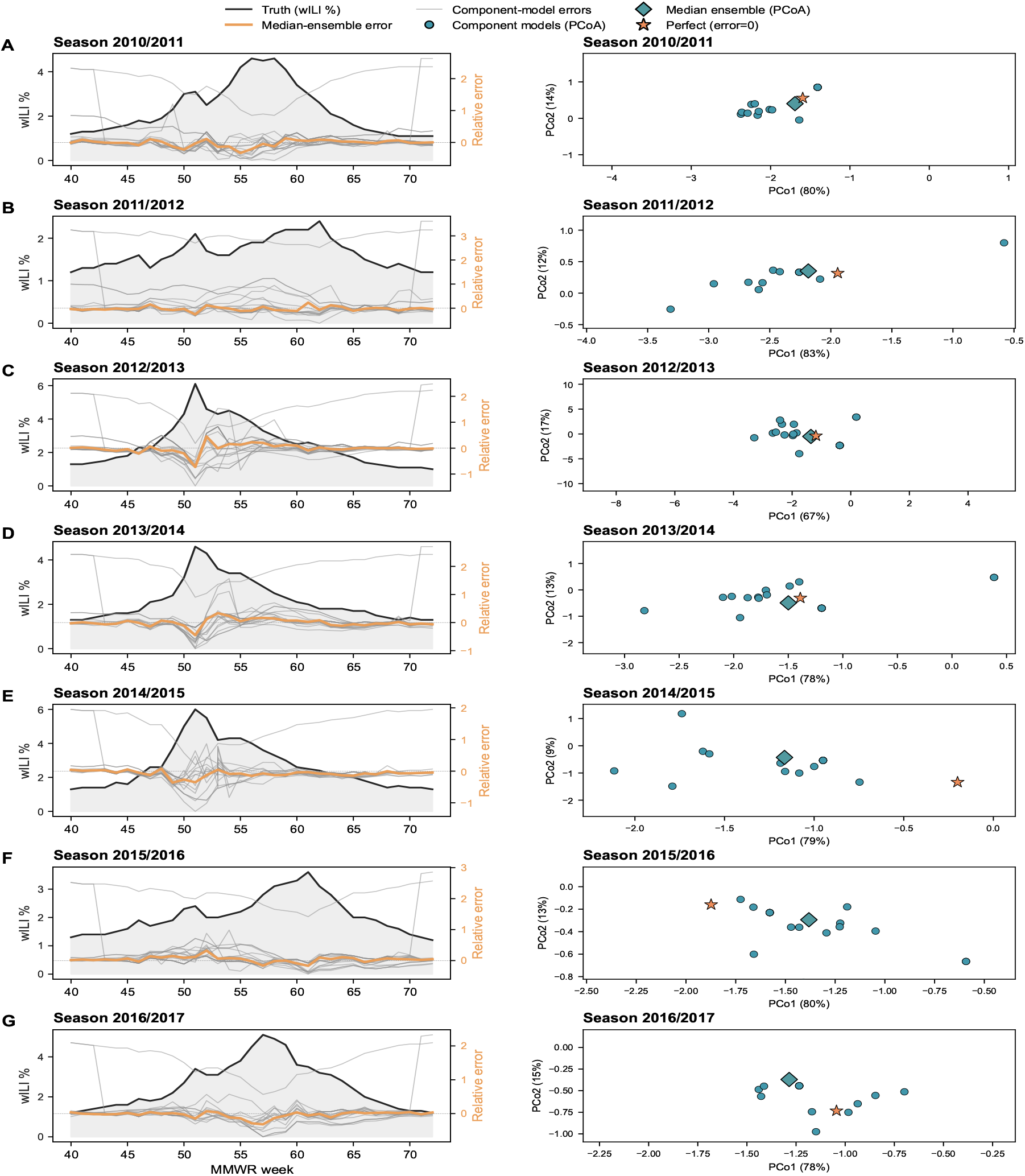
Structural errors in real-time forecasts for influenza ILI at *h* = 0. Per-season component-model error trajectories (grey) overlaid on weighted ILI percentage (wILI%, black) and the median-ensemble error trajectory (red) at left; classical PCoA on Euclidean distances of the per-model error time series at right, with the median ensemble (orange diamond) and a synthetic “Perfect” (red star) point shown for reference. Seven panel pairs, one per ILI season (2010–11 through 2016–17). The component-model cluster is tight and the “PERFECT” point separates cleanly along PCo1 in every season. For legibility, each PCoA panel is zoomed to the component-model cluster, so a few extreme-error models (e.g., the uniform baseline) fall outside the plotted range in some seasons; all component models are retained in the analysis. Left-axis trajectories are relative errors, normalized by the season-mean wILI.

**Figure S2:**
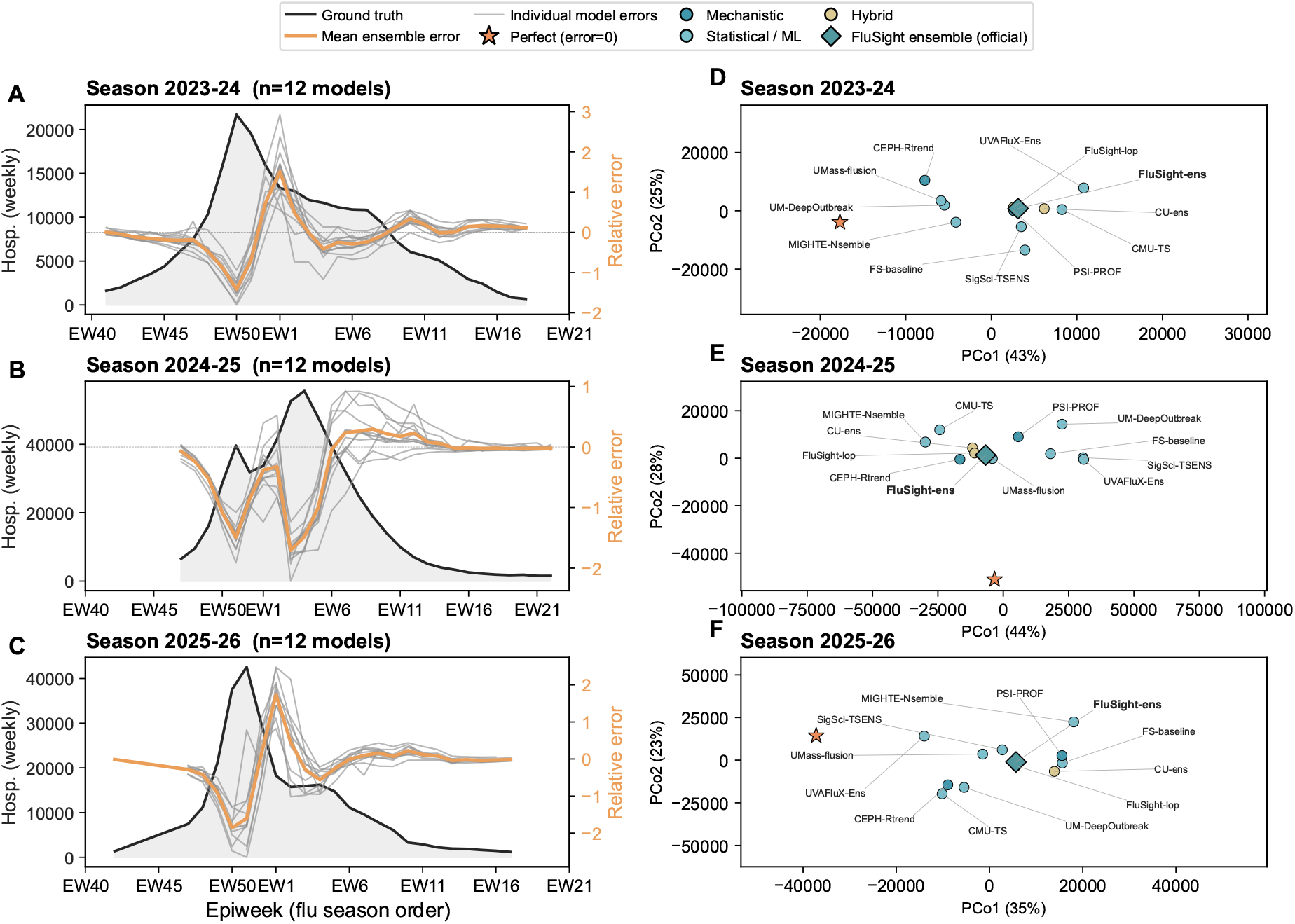
Structural errors in real-time forecasts for influenza hospitalizations at *h* = 2. (A-C) 2-week-ahead forecast errors of component models (gray solid lines) and the unweighted ensemble (orange solid lines), overlaid with the weekly hospitalizations at the national level (black solid lines). Forecast errors in (A–C) are shown as relative errors, each normalized by that season’s mean target value (right axis). We include 12 models with no missing forecasts submitted to the CDC FluSight challenge. For the 2025-2026 season, data are shown up to Epiweek 18 in 2026, when the analysis was performed. (D-F) Two-dimensional Principal Coordinates Analysis (PCoA, classical metric multidimensional scaling) of 2-week-ahead forecast-error time series, computed on pairwise Euclidean distances, with a synthetic “Perfect” (zero-error) reference included for context. Component model structures are grouped into mechanistic, statistical/ML, and hybrid approaches, shown in different colors. The official FluSight ensemble is shown using teal diamonds. Coral stars represent the embedding of perfect forecasts as a reference. For legibility, each PCoA panel is zoomed to the component-model cluster, so a few extreme-error models (e.g., the uniform baseline) fall outside the plotted range in some seasons; all component models are retained in the analysis.

**Figure S3:**
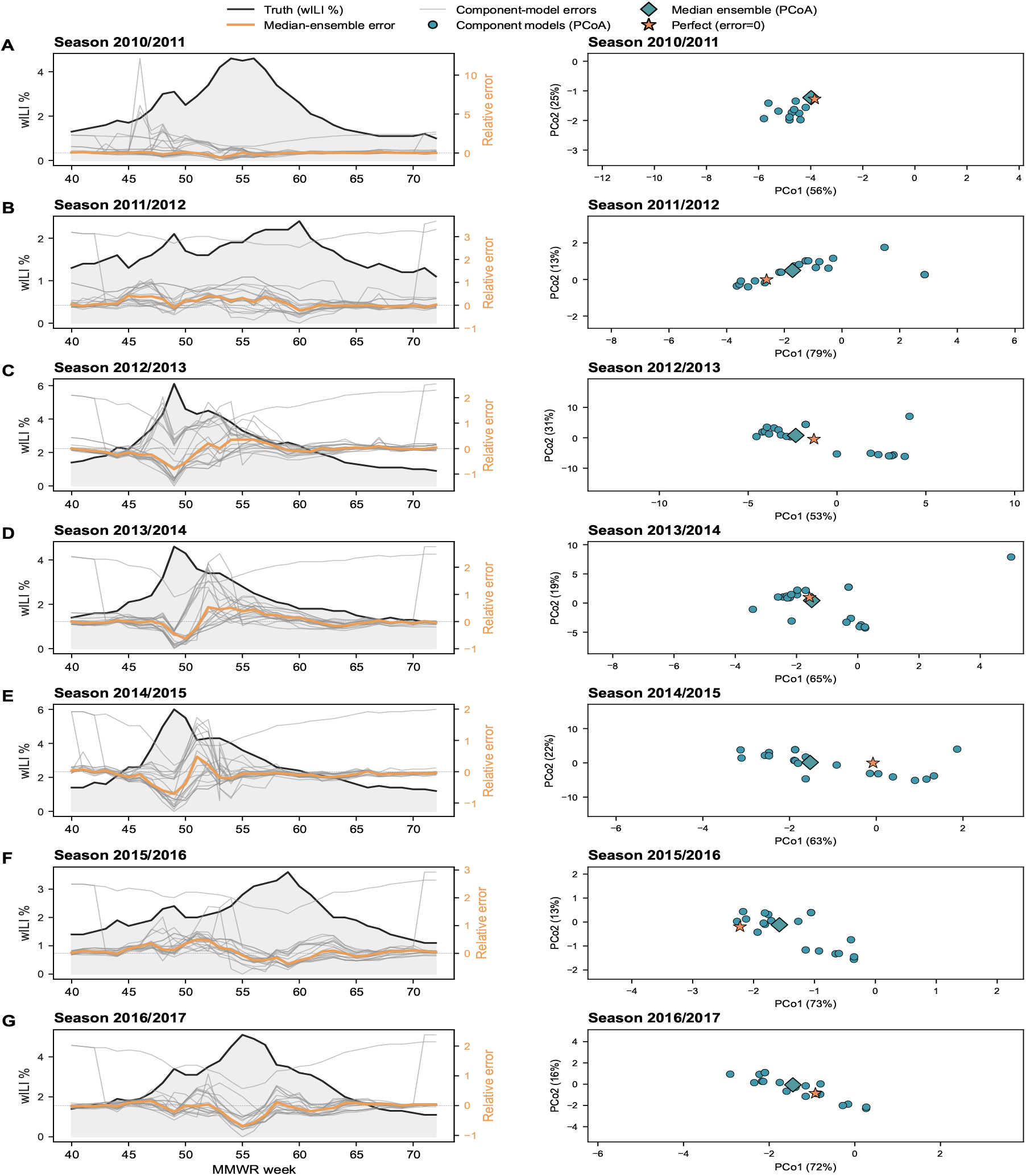
Structural errors in real-time forecasts for influenza ILI at *h* = 2. Per-season component-model error trajectories (grey) overlaid on weighted ILI percentage (wILI%, black) and the median-ensemble error trajectory (red) at left; classical PCoA on Euclidean distances of the per-model error time series at right, with the median ensemble (orange diamond) and a synthetic “Perfect” (red star) point shown for reference. Seven panel pairs, one per ILI season (2010–11 through 2016–17). The component-model cluster is tight and the “PERFECT” point separates cleanly along PCo1 in every season. For legibility, each PCoA panel is zoomed to the component-model cluster, so a few extreme-error models (e.g., the uniform baseline) fall outside the plotted range in some seasons; all component models are retained in the analysis. Left-axis trajectories are relative errors, normalized by the season-mean wILI.

## STS-Residual implementation details

### Common input features

For each (location, week) prediction point, both deep branches use the same *D*-dimensional per-step feature vector: the relative residual (*ε*) of each individual component model, the relative residual of the training ensemble, and a sine/cosine encoding of the epiweek. The two branches differ only in *which* length-*W* residual window they read, dictated by how each is trained: the per-horizon LSTM reads the leak-free nowcast (*h*=0) window for every target horizon, while the single jointly-trained GAT reads the horizon-specific residuals *ε*^*h*^ from a window shifted back by *h*+1 issue weeks (so the most recent target stays at week *t* − 1). For hospitalizations the FluSight-ensemble is excluded from the per-model features (it serves as the evaluation benchmark, though it is one of the 12 retained models), leaving 11 model residuals; for ILI the constant-weight benchmark is a separate weighted aggregate rather than one of the 21 component models, so all 21 are used. This gives *D* = 14 for hospitalizations (11 model residuals + 1 ensemble + 2 epiweek) and *D* = 24 for ILI (21 model residuals + 1 ensemble + 2 epiweek). The window length *W* = 4 weeks was selected from *{*3, 4, 6, 8*}* by inner cross-validation on the training fold.

### Seasonal branch

The seasonal branch produces a per-(location, horizon) correction

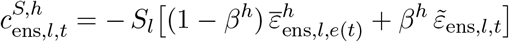

where 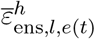 is the average ensemble residual at the same epiweek and horizon across training-fold seasons, and 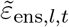 is the mean of the most recent *L* horizon-0 residuals. The mixing coefficient *β* is selected per horizon by inner LOSO cross-validation on the training fold from the grid *β* ∈ *{*0, 0.05, 0.10, …, 1.00*}*.

### Temporal branch (LSTM)

A single-layer LSTM with hidden dimension 32 reads the (*W* = 4) *× D* feature window. The hidden state at the final step, *h*_*T*_ ∈ ℝ ^32^, is passed through a two-layer MLP (32 → 16 → 1, ReLU + dropout) to produce the scalar correction *c*_*T*_. Architecture sweep (hidden ∈ {32, 64}, layers ∈ {1, 2}, *W* ∈ {3, 4, 6, 8}, dropout ∈ {0.1, 0.2}) is performed by inner LOSO cross-validation on the training fold; the configuration above was selected per horizon by inner-fold MAE. Training: 300 epochs of Adam (learning rate 10^−3^, weight decay 10^−5^, batch size 256) with a ReduceLROnPlateau scheduler (patience 20, factor 0.5) and a mean-squared-error loss on the relative residual target. **A separate LSTM is trained per horizon**.

#### Spatial branch (LSTM + GAT)

Each location’s (*W × D*) window is first encoded by a shared single-layer LSTM (hidden 32; weights tied across locations) to give a per-node 32-dimensional embedding. A single graph-attention layer with 4 attention heads (32-dim output, dropout 0.1) operates on a binary adjacency mask: nodes are locations and edges are geographic-adjacency relations between states (with self-loops on every node). The post-GAT 32-dimensional per-node embedding is passed through the same two-layer MLP head (32 → 16 → 1) to produce *c*_*G*_ for that location. Three states with no land neighbors – Alaska, Hawaii, and Puerto Rico – are treated as isolated nodes (self-loop only). Because each LOSO fold yields only ≈55 full-graph snapshots per horizon, **a single GAT is trained jointly across all four horizons**; unlike the temporal branch it reads the horizon-specific lagged residuals *ε*^*h*^ (from the window shifted back by *h*+1 issue weeks), which lets the shared model distinguish the four lead times. Training uses the same Adam settings as the LSTM branch.

#### Adaptive per-horizon blend

The three branch corrections are combined with a per-horizon convex weighting

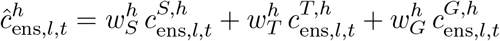

The weight triple 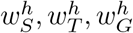 is selected separately for each horizon *h* ∈ *{*0, 1, 2, 3*}* by inner leave-one-season-out cross-validation on the training fold, searching the simplex grid with increment 0.05. The resulting weights are reported in Table S3.

**Table S3:** Mean adaptive blend weights (*w*_*S*_, *w*_*T*_, *w*_*G*_) selected by inner-CV per horizon, averaged across LOSO training folds. For hospitalizations, the seasonal branch dominates at every horizon, with the temporal and spatial branches contributing modest shares that grow at longer lead times; for ILI, the seasonal branch dominates at every horizon, with the temporal and spatial branches contributing only small shares.

| Horizon | hospitalization |  |  | ILI |  |  |
| --- | --- | --- | --- | --- | --- | --- |
| | $w_S$ | $w_T$ | $w_G$ | $w_S$ | $w_T$ | $w_G$ |
| $h = 0$ | 0.80 | 0.14 | 0.06 | 0.83 | 0.06 | 0.11 |
| $h = 1$ | 0.60 | 0.10 | 0.30 | 0.59 | 0.16 | 0.26 |
| $h = 2$ | 0.71 | 0.04 | 0.26 | 0.54 | 0.16 | 0.30 |
| $h = 3$ | 0.70 | 0.30 | 0.00 | 0.66 | 0.14 | 0.20 |

#### Applying the correction

Before the adaptive blend, each branch’s normalized correction *c*_*r*_*/S*_*l*_ is clipped to [−1, 1] (an overcorrection safeguard used throughout). At inference time, the resulting scalar 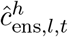 is added uniformly to every predictive quantile *q*_*τ*_ of the official ensemble distribution; the shifted quantiles are clipped at zero and monotonized 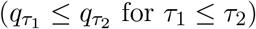.

#### Cross-validation protocol

All hyperparameters (lookback length *W*, seasonal mix *β*, LSTM architecture, blend weights) are selected per-horizon by inner LOSO cross-validation on the outer training fold only; the held-out test season is never seen during selection. The outer loop holds out each season in turn (3 folds for hospitalizations, 7 folds for ILI).

### Modeling setups and evaluation targets

STS-Residual was evaluated under two related hospitalization setups, each designed to answer a different operational question. In both, the framework was used as a post-prediction correction layer that learned a single correction from historical forecasts—the residual of the equal-weight mean of the 12 retained models—and applied it to a pre-existing ensemble forecast. The two setups share the same learning target and the same input features; they differ only in the ensemble to which the correction is applied and against which it is benchmarked: the official FluSight-ensemble (primary) or the same mean ensemble (secondary). The setups are summarized in Table S4.

#### Main hospitalization analysis

The primary hospitalization analysis evaluated whether STS-Residual could improve the official CDC FluSight-ensemble for weekly influenza hospital admissions. The supervised learning target was the residual of the equal-weight, quantile-wise mean of the 12 retained models, normalized by the location scale *S*_*l*_. The temporal and spatial branches used as input features the residuals of the 11 retained submissions other than the FluSight-ensemble, together with this mean-ensemble residual and a cyclic epiweek encoding; for each forecast point, only lagged residuals whose target observations would have been available before the correction time were used. The learned correction was then applied to the official FluSight-ensemble forecast issued for the same location, week, and horizon, and the corrected forecast was evaluated against the uncorrected FluSight-ensemble. The FluSight-ensemble itself is therefore never used as an input feature; it serves only as the forecast that is corrected and as the evaluation benchmark.

**Table S4:** Modeling setups used for STS-Residual evaluation. Both hospitalization setups share the same training target (the normalized residual of the equal-weight, quantile-wise mean of the 12 retained models) and the same input features (the residuals of the 11 retained submissions other than the FluSight-ensemble, the mean-ensemble residual, and a cyclic epiweek encoding). They differ only in the ensemble to which the learned correction is applied and against which it is benchmarked.

| Analysis | Residual learned (training target) | Correction applied to & benchmark |
| --- | --- | --- |
| Main (primary) | Equal-weight mean of the 12 retained models | Official FluSight-ensemble |
| Secondary | Equal-weight mean of the 12 retained models | Equal-weight mean of the 12 retained models |

#### Retained hospitalization analysis

We performed a secondary hospitalization analysis using only the retained forecast submissions. In this setup, the base forecast was the equal-weight, quantile-wise mean of the retained hospitalization submissions. The residual target was the normalized residual of this retained-model mean ensemble, and the learned correction was applied back to the same retained-model mean ensemble. The corrected forecasts were evaluated against the uncorrected retained-model mean ensemble. This analysis tests whether performance gains arise from learnable residual structure within the retained forecast pool.

#### ILI analysis

For ILI, the constant-weight (CW) ensemble distributed with the FluSight Network archive serves as *both* the training ensemble (whose residual is learned) and the official benchmark. The two hospitalization setups above therefore coincide for ILI, and a single CW-based comparison is reported throughout.

### Example corrected forecasts

Figure S4 shows example national-level corrected forecasts for representative hospitalization and ILI seasons; per-horizon performance metrics for all locations are reported in the main-text performance table (Table 1).

**Figure S4:**
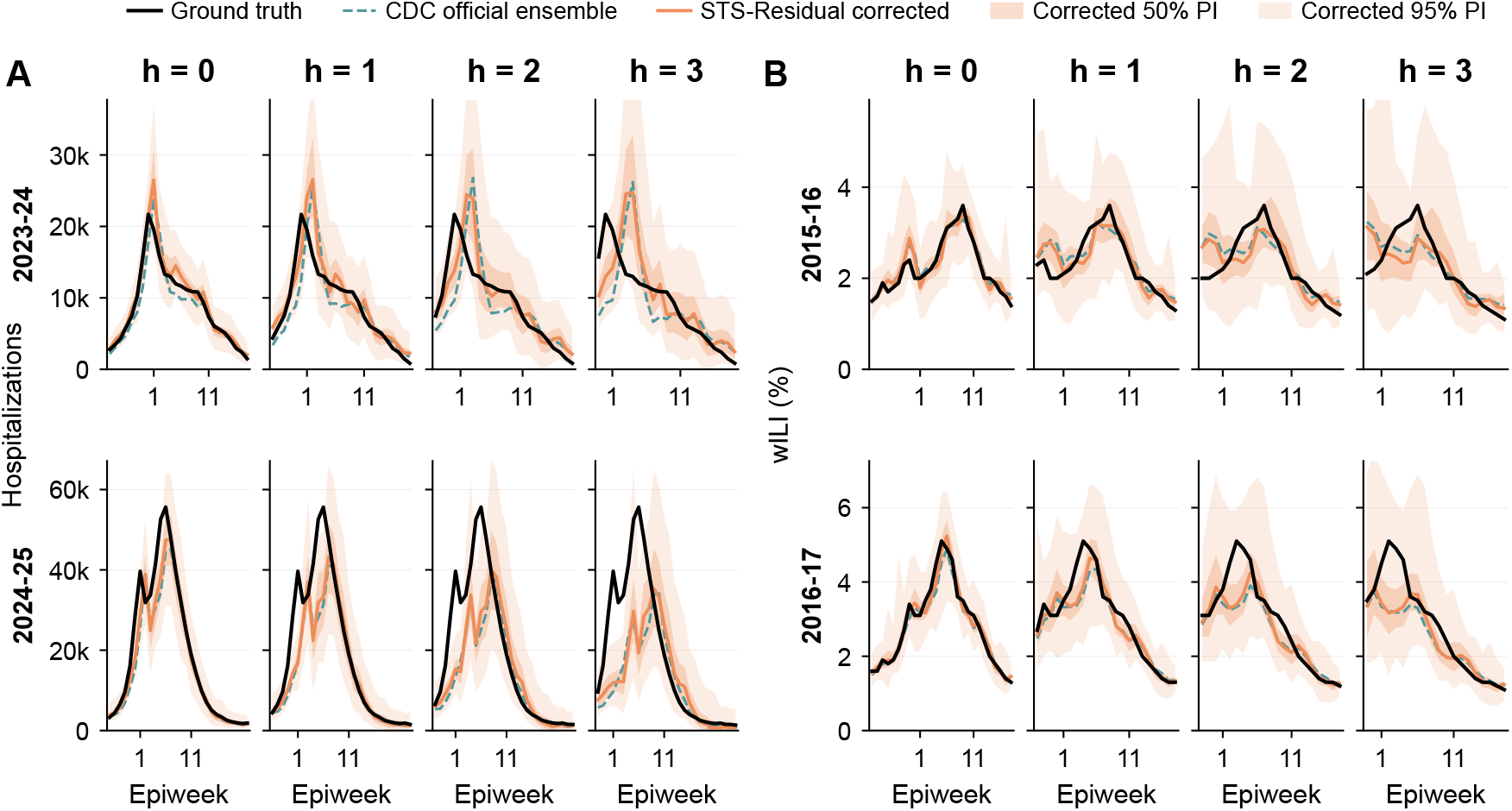
Example corrected national forecasts. (A) 0-to 3-week-ahead influenza hospitalization forecasts for the 2023–2024 and 2024–2025 seasons; (B) 0-to 3-week-ahead ILI forecasts for the 2015–2016 and 2016–2017 seasons. Ground truth (black), original ensemble (teal dashed), and STS-Residual corrected forecasts (solid coral) are compared; shaded coral bands show the corrected 50% (darker) and 95% (lighter) prediction intervals (shift-only).

### Correction relative to the equal-weight mean ensemble

The hospitalization results in the main text benchmark the STS-Residual–corrected forecast against the CDC FluSight-ensemble, which aggregates a broader set of submissions than the 12 component models retained here. To confirm that the improvement is not merely an artifact of this difference in model composition, we repeated the hospitalization evaluation using the equal-weight, quantile-wise mean of the 12 selected models as *both* the training ensemble and the benchmark: the learned correction is applied to this mean ensemble and scored against it. STS-Residual remains consistently beneficial (Table S5), reducing WIS by 10.1%, 5.5%, 3.5%, and 1.9% pooled across the 52 locations (and by 17.9%, 10.5%, 8.2%, and 4.3% at the US national level) for *h* = 0 through *h* = 3. These reductions are slightly smaller than those obtained against the FluSight-ensemble (main-text Table 1) but follow the same pattern across horizons, indicating that the correction captures genuine residual structure rather than exploiting the difference between the selected pool and the official ensemble.

**Table S5:** Secondary hospitalization benchmark: STS-Residual improvement (%) when the correction is applied to, and evaluated against, the equal-weight quantile-wise mean of the 12 selected models, by horizon. Sub-national = all 52 locations; National = US row only.

| Scope | Metric | $h=0$ | $h=1$ | $h=2$ | $h=3$ |
| --- | --- | --- | --- | --- | --- |
| Sub-national (n=52) | WIS imp. % | 10.1 | 5.5 | 3.5 | 1.9 |
|  | MAE imp. % | 9.5 | 5.3 | 2.8 | 1.5 |
| US National | WIS imp. % | 17.9 | 10.5 | 8.2 | 4.3 |
|  | MAE imp. % | 22.0 | 11.4 | 7.7 | 4.6 |

### Observed epidemic-state input features

The temporal and spatial branches in the main analysis use residual-based features only: the per-model and training-ensemble residuals together with a cyclic epiweek encoding. During development we also considered feeding both learners two *observed-state* signals—the normalized observed level *y*_*l,s*_*/S*_*l*_ and its week-over-week change (*y*_*l,s*_ − *y*_*l,s*−1_)*/S*_*l*_—which give the model direct access to the current epidemic level and its trend. Both are computed from past observations only (the change uses the previous week’s value), so they remain leak-free under the same one-week reporting-delay assumption used for the residual features.

Adding the observed-state features yielded similar overall performance (Table S6), with only small changes across horizons and no consistent gain. The main analysis therefore retains the residual-only features for simplicity.

**Table S6:** Effect of adding observed epidemic-state features. Both the LSTM and GAT branches additionally receive the normalized observed level and its week-over-week change; all other settings match the main analysis. Improvements (%) are relative to the FluSight-ensemble, by horizon. “Main” is the residual-only configuration used throughout the paper.

| Scope / metric | Configuration | $h=0$ | $h=1$ | $h=2$ | $h=3$ |
| --- | --- | --- | --- | --- | --- |
| Sub-national (52) WIS | Main | 11.6 | 6.9 | 5.0 | 2.7 |
|  | + observed state | 10.9 | 7.2 | 4.7 | 2.5 |
| Sub-national (52) MAE | Main | 10.3 | 6.0 | 3.9 | 2.0 |
|  | + observed state | 9.7 | 6.7 | 3.5 | 2.0 |
| US National WIS | Main | 19.9 | 12.5 | 9.9 | 4.8 |
|  | + observed state | 19.1 | 12.6 | 7.7 | 4.3 |
| US National MAE | Main | 22.3 | 13.1 | 8.6 | 5.3 |
|  | + observed state | 20.7 | 15.3 | 5.9 | 5.0 |

### Per-state and per-region performance breakdown

Tables S7 and S8 report the MAE-improvement (%) of STS-Residual relative to the official ensemble for every individual location, by horizon. Hospitalization states are sorted by the row mean across the four horizons. Negative values indicate locations where the correction degrades MAE relative to the benchmark; these are concentrated in low-burden or isolated locations (PR, HI, AK).

**Table S7:** Hospitalization: per-location MAE improvement (%) of STS-Residual vs. FluSight-ensemble. States are sorted by the row-mean across *h* = 0, 1, 2, 3. Negative values are losses.

| State | $h=0$ | $h=1$ | $h=2$ | $h=3$ | Mean | State | $h=0$ | $h=1$ | $h=2$ | $h=3$ | Mean |
| --- | --- | --- | --- | --- | --- | --- | --- | --- | --- | --- | --- |
| US | 22.3 | 13.1 | 8.6 | 5.3 | 12.3 | UT | 6.6 | 3.9 | 5.7 | 2.7 | 4.7 |
| MO | 15.6 | 9.3 | 10.5 | 7.3 | 10.7 | ME | 4.0 | 5.0 | 4.8 | 4.9 | 4.6 |
| TX | 24.2 | 11.2 | 5.2 | 0.5 | 10.3 | LA | 10.4 | 4.6 | 1.7 | 1.6 | 4.6 |
| IN | 18.6 | 10.6 | 6.4 | 4.6 | 10.1 | MD | 10.5 | 1.1 | 4.0 | 2.6 | 4.5 |
| MI | 15.0 | 6.6 | 8.9 | 4.7 | 8.8 | NC | 9.3 | 4.9 | 2.0 | 1.2 | 4.3 |
| IA | 14.4 | 10.0 | 6.4 | 3.3 | 8.5 | RI | 5.4 | 6.0 | 2.9 | 2.7 | 4.2 |
| ID | 12.7 | 6.1 | 10.5 | 3.9 | 8.3 | MS | 9.4 | 4.4 | 2.2 | 0.5 | 4.1 |
| KY | 11.2 | 8.7 | 8.8 | 3.6 | 8.1 | MT | 6.2 | 3.5 | 2.8 | 3.2 | 3.9 |
| NY | 14.4 | 8.0 | 5.3 | 4.3 | 8.0 | AZ | 6.9 | 3.2 | 3.1 | 2.2 | 3.8 |
| MA | 15.7 | 8.1 | 4.8 | 2.4 | 7.7 | NM | 1.7 | 6.2 | 4.8 | 2.1 | 3.7 |
| OR | 9.9 | 4.9 | 9.2 | 6.6 | 7.7 | GA | 10.2 | 4.8 | 1.6 | -2.4 | 3.6 |
| WI | 10.1 | 9.9 | 4.7 | 2.6 | 6.8 | WV | 3.1 | 4.5 | 3.8 | 2.6 | 3.5 |
| NE | 10.2 | 7.3 | 5.1 | 3.3 | 6.5 | SD | 3.1 | 2.5 | 3.0 | 5.3 | 3.5 |
| NJ | 12.4 | 7.0 | 3.3 | 2.8 | 6.4 | DE | 3.3 | 4.2 | 3.4 | 1.3 | 3.1 |
| IL | 6.3 | 8.8 | 6.1 | 3.3 | 6.2 | VA | 2.3 | -1.1 | 5.2 | 5.2 | 2.9 |
| AR | 8.4 | 7.9 | 5.0 | 2.9 | 6.0 | NH | 3.7 | 4.0 | 0.4 | 2.6 | 2.7 |
| KS | 7.2 | 7.4 | 5.8 | 2.7 | 5.8 | ND | 3.1 | 3.4 | 1.2 | 1.1 | 2.2 |
| AL | 14.8 | 6.4 | 2.9 | -1.0 | 5.8 | VT | -1.2 | -1.1 | 4.6 | 3.1 | 1.4 |
| TN | 7.3 | 5.5 | 7.3 | 2.7 | 5.7 | WY | 0.5 | 1.3 | 3.0 | -0.3 | 1.1 |
| CO | 6.5 | 10.8 | 2.3 | 2.7 | 5.6 | DC | 1.3 | 0.8 | 1.8 | -0.1 | 1.0 |
| OK | 11.1 | 4.2 | 3.8 | 2.3 | 5.3 | NV | -2.6 | 3.7 | 0.5 | 1.3 | 0.7 |
| OH | 9.7 | 6.3 | 3.7 | 1.5 | 5.3 | CA | 8.8 | 1.1 | -3.9 | -3.4 | 0.6 |
| FL | 10.8 | 6.7 | 1.2 | 2.1 | 5.2 | WA | -10.2 | 4.9 | 3.4 | 1.3 | -0.2 |
| CT | 7.4 | 6.3 | 3.6 | 3.2 | 5.1 | PR | -1.7 | -5.0 | 0.0 | -2.0 | -2.2 |
| PA | 8.6 | 6.4 | 4.0 | 1.3 | 5.1 | AK | -6.1 | -5.9 | -1.9 | -2.0 | -4.0 |
| MN | 4.9 | 4.4 | 6.4 | 4.5 | 5.1 | HI | -4.6 | -9.4 | -2.7 | -12.1 | -7.2 |
| SC | 11.4 | 6.2 | 3.0 | -0.4 | 5.0 |  |  |  |  |  |  |
| **Mean (53 locations):** $h = 0$: 7.6 $h = 1$: 5.0 $h = 2$: 4.0 $h = 3$: 2.0 Overall: 4.7 | | | | | | | | | | | |

### Ablation and sensitivity analyses

We ablate the three STS-Residual branches by re-running the leave-one-season-out blend inner-CV with restricted branch subsets, holding the already-trained per-branch corrections (Seasonal *c*_*S*_, Temporal *c*_*T*_, Spatial *c*_*G*_) fixed. Each ablation thus measures the marginal value of including or excluding a branch from the adaptive blend; the deep models themselves are not retrained, ensuring the ablation isolates the contribution of each branch rather than confounding it with optimization variance.

Seven configurations are evaluated against the official ensemble for both datasets (Table S9):

- **S only / T only / G only**: a single branch is used as the correction (*w* fixed at 1 for that branch).
- **S+T / S+G / T+G**: two-branch convex blend with the optimal mix selected by inner-CV from the 0.05 grid.
- **Full (paper)**: the three-branch adaptive blend used throughout the main paper.

The branch contributions differ between datasets. For **hospitalizations** the **seasonal branch** accounts for most of the improvement, leading at the nowcast and one-week horizons (*S* alone 14.9% and 9.6%), while the **temporal and spatial branches** add modest gains at the two- and three-week horizons; the full blend gives the best mean improvement (8.8%), only marginally above the seasonal branch alone (7.8%). For **ILI** the **Seasonal branch accounts for essentially the entire correction**: *S* alone (mean 2.8%) and any seasonal-containing pair (*S*+*T* 3.5%, *S*+*G* 3.4%) match the full blend (3.4%), whereas the Temporal and Spatial branches are weak or harmful on their own (mean −0.3% and v3.2%). On the coarse 11-region ILI graph the temporal and spatial branches carry little independent signal, and the adaptive blend down-weights them.

**Table S8:** ILI: per-region MAE improvement (%) of STS-Residual vs. the constant-weight ensemble. Improvements are largest at the nowcast horizon and decline with lead time, turning negative at longer horizons in several regions.

| Region | $h=0$ | $h=1$ | $h=2$ | $h=3$ | Mean |
| --- | --- | --- | --- | --- | --- |
| HHS Region 9 | 11.8 | 8.0 | 7.5 | 7.2 | 8.6 |
| HHS Region 10 | 5.2 | 9.7 | 7.8 | 9.7 | 8.1 |
| HHS Region 8 | 4.9 | 8.5 | 9.3 | 3.7 | 6.6 |
| HHS Region 6 | 11.2 | 6.3 | 2.8 | -0.2 | 5.0 |
| US National | 8.5 | 10.8 | 3.3 | -3.9 | 4.7 |
| HHS Region 2 | 10.3 | 5.0 | 2.9 | -0.7 | 4.4 |
| HHS Region 4 | 4.8 | 6.5 | 1.6 | -0.1 | 3.2 |
| HHS Region 7 | 6.9 | 3.6 | 1.5 | -3.5 | 2.1 |
| HHS Region 3 | 4.0 | -1.2 | -7.6 | -8.6 | -3.3 |
| HHS Region 5 | 3.9 | -2.3 | -5.7 | -10.8 | -3.7 |
| HHS Region 1 | 9.6 | -5.9 | -9.2 | -11.7 | -4.3 |
| **Mean (11)** | **7.4** | **4.5** | **1.3** | **-1.7** | **2.8** |

#### Interpretation

The ablation makes the role of the adaptive blend explicit: the per-horizon weighting routes weight to whichever branch carries signal, so the same framework adapts to two very different datasets without manual tuning. On the rich 53-location hospitalization data it leans on the Seasonal branch at the shortest horizons and the Temporal and Spatial branches at longer horizons, and the full blend gives the best mean improvement (8.8%), with the strongest two-branch combination (*S*+*T*, 8.2%) trailing by less than 1 pp. On the coarse 11-region ILI data it leans almost entirely on the Seasonal branch, and any seasonal-containing configuration matches the full blend to within ~0.1 pp. Carrying all three branches therefore costs little while guaranteeing the informative branch is always present.

#### Sensitivity to *β* (Seasonal mix)

The blending coefficient *β* between the epiweek-mean look-up and the recent-error correction 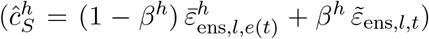 is selected per-horizon by inner-CV on a 0.05 grid. Across LOSO folds, the selected *β* has median 0.60 for hospitalization and 0.60–0.70 for ILI at *h* = 0, decreasing towards 0 at *h* ≥ 2, consistent with the intuition that the recent-error signal is most informative at the nowcast horizon.

### Robustness to changing component-model composition

We probe whether STS-Residual’s gains depend on the exact composition of the component-model pool with a transfer Monte-Carlo experiment on the hospitalization target. At each of 10 iterations, we draw an independent random subset of 7 of the 9 individual-team models *separately for every season*, rebuild that season’s equal-weight quantile ensemble from its own subset, and train STS-Residual under the same leave-one-season-out protocol with identity-agnostic inputs (the ensemble residual and a cyclic epiweek encoding only). The corrector is then applied to the held-out season’s subset ensemble—whose composition was never seen during training—and scored against that same uncorrected subset ensemble. Because rebuilt subset ensembles are occasionally unstable, each branch’s normalized correction is capped to [−1, 1] before the adaptive blend (the same overcorrection safeguard used in the main analysis).

**Table S9:** Ablation: STS-Residual MAE improvement (%) relative to the official benchmark ensemble, per horizon, when only a subset of the three branches is allowed to contribute to the blend. Within each dataset, the best configuration at each horizon is bolded. For hospitalizations, the full blend gives the best mean improvement, with the temporal and spatial branches contributing more at the longer horizons; for ILI, the seasonal branch alone accounts for essentially all of the improvement, and adding the temporal or spatial branch neither helps nor hurts.

| Dataset | Configuration | $h = 0$ | $h = 1$ | $h = 2$ | $h = 3$ | Mean |
| --- | --- | --- | --- | --- | --- | --- |
| hospitalization | Seasonal only ($S$) | 14.9 | 9.6 | 4.3 | 2.5 | 7.8 |
| | Temporal only ($T$) | 3.7 | 7.5 | 6.5 | 6.4 | 6.0 |
| | Spatial only ($G$) | 0.8 | 4.6 | 6.8 | 3.0 | 3.8 |
| | $S + T$ | 13.4 | 9.4 | 4.6 | 5.6 | 8.2 |
| | $S + G$ | 14.2 | 9.4 | 5.7 | 2.7 | 8.0 |
| | $T + G$ | 2.8 | 7.7 | 7.5 | 5.9 | 6.0 |
|  | **Full ($S+T+G$, paper)** | 14.1 | 9.3 | 6.2 | 5.5 | **8.8** |
| ILI | Seasonal only ($S$) | 6.9 | 4.4 | 0.9 | -1.0 | 2.8 |
| | Temporal only ($T$) | 5.9 | 1.5 | -1.9 | -6.6 | -0.3 |
| | Spatial only ($G$) | -0.1 | -1.2 | -3.2 | -8.2 | -3.2 |
| | $S + T$ | **8.4** | 5.1 | 1.5 | -0.9 | **3.5** |
| | $S + G$ | 7.8 | **5.3** | **2.0** | -1.6 | 3.4 |
| | $T + G$ | 5.9 | 1.5 | -1.3 | -5.8 | 0.1 |
|  | **Full ($S+T+G$, paper)** | 7.9 | 4.9 | 1.7 | -1.1 | 3.4 |

Across the 3 *×* 10 = 30 per-season runs (Table S10), the safeguarded correction improves on the subset-rebuilt ensemble at every horizon, with mean WIS reductions of 12.1%, 7.6%, 7.3%, and 4.6% (MAE reductions of 14.6%, 7.0%, 7.1%, and 4.6%) and a 90–100% win rate. The consistently positive safeguarded transfer indicates that the learned residual structure is genuinely shared across ensembles built from different model sets rather than tied to specific training-time identities.

**Table S10:**
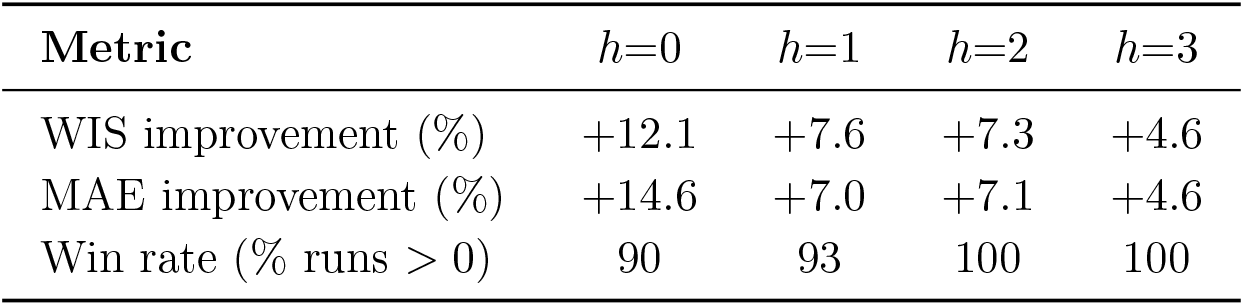
Transfer robustness (hospitalization): STS-Residual trained on a random 7-of-9 subset drawn independently per season, applied to the held-out season’s subset ensemble and scored against that same uncorrected ensemble. Improvements (%) are location-averaged over the 3 *×* 10 = 30 runs.

| Metric | $h=0$ | $h=1$ | $h=2$ | $h=3$ |
| --- | --- | --- | --- | --- |
| WIS improvement (%) | +12.1 | +7.6 | +7.3 | +4.6 |
| MAE improvement (%) | +14.6 | +7.0 | +7.1 | +4.6 |
| Win rate (% runs $> 0$) | 90 | 93 | 100 | 100 |

### Robustness to real-time surveillance revisions

Confirmed influenza admissions reported through NHSN are subject to retrospective revision as hospitals correct reporting errors and submit backfilled data [40], so a forecast issued in real time sees only the provisional observations available at the forecast date. We test whether STS-Residual’s gains survive this for the two hospitalization seasons (2024–25 and 2025–26) for which both first-released and finalized weekly values are available from the Delphi Epidata API [47]. Holding out one NHSN season at a time, we recomputed that season’s lagged residual features against its *first-released* observations (the training seasons retained their finalized values) and applied the same [− 1, 1] overcorrection safeguard. The corrected forecast was then scored two ways: against the first-released targets (the truth available in real time) and against the finalized targets (the eventually-revised truth).

Pooled across both seasons and all 53 locations (Table S11), the corrected ensemble improves on the FluSight-ensemble at every horizon under *both* scoring conventions. Provisional and finalized admissions differ on 74% of the scored location-weeks, yet the two sets of improvements agree to within 0.5 percentage points, indicating that the gain is insensitive to whether later data revisions are taken into account; only the longest horizon shows the expected mild attenuation.

**Table S11:** Real-time robustness (hospitalization): test-season residuals computed against first-released NHSN admissions, with the corrected forecast scored against both the first-released (real-time) and the finalized targets. Pooled over the two NHSN seasons (2024–25, 2025–26) and all 53 locations, relative to the FluSight-ensemble.

| Scoring truth | Metric | $h=0$ | $h=1$ | $h=2$ | $h=3$ |
| --- | --- | --- | --- | --- | --- |
| First-released | WIS imp. % | +12.6 | +9.6 | +4.7 | +1.6 |
|  | MAE imp. % | +11.5 | +11.4 | +3.0 | +0.9 |
| Finalized | WIS imp. % | +13.1 | +9.2 | +4.7 | +1.7 |
|  | MAE imp. % | +11.3 | +9.8 | +3.5 | +1.0 |

